# Socioeconomic and temporal heterogeneity in SARS-CoV-2 exposure and disease in England from May 2020 to February 2023

**DOI:** 10.1101/2024.11.11.24317098

**Authors:** Christian Morgenstern, Thomas Rawson, Wes Hinsley, Pablo N. Perez Guzman, Samir Bhatt, Neil M. Ferguson

**Author notes:** Contributed equally (Samir Bhatt, Neil M. Ferguson).

## Abstract

The impact of COVID-19 varied significantly by deprivation, ethnicity, and policy measures. We analysed individual-level data on 12,310,485 first SARS-CoV-2 Pillar-2-PCR-confirmed infections, 439,083 hospitalisations, 107,823 deaths, and vaccination records in England from May 2020 to February 2022. Poisson regression models adjusted for demographic and temporal factors showed higher incidence rate ratios (IRRs) for severe outcomes in the most deprived areas compared to the least. We note higher IRRs for severe outcomes for all non-White relative to White ethnicities. The magnitude of IRRs for both deprivation and ethnicities declined from the wild-type to the omicron periods for severe outcomes. For infections, we observed IRRs above one for non-White ethnicities during the wild-type and alpha periods. Vaccination significantly reduced risks across all groups. For severe outcomes, pre-existing health inequalities led to large and persistent disparities. For infections, measures must be structured with ethnicity and deprivation in mind early in a pandemic.

## Introduction

By December 2023, the COVID-19 pandemic had caused over 20.5 million confirmed cases and over 175,000 deaths in England **(*1, 2*)**. The pandemic did not impact individuals equally; individual-based studies have explored the links between deprivation, ethnicity, and other factors on health outcomes to varying levels **(*3*–*8*)**.

Past studies have explored heterogeneity in risk in England **(*9, 10*)** and Scotland **(*11*)** using the Index of Multiple Deprivation (IMD) **(*12*)** at the Lower-Tier Local Authority (LTLA) level. LTLAs are areas where local government provides services, and the UK Office of National Statistics (ONS) provides an extensive set of data, including population age distribution, ethnicity, and measures of deprivation. These measures vary substantially across LTLAs, as did the number of PCR-confirmed infections, hospitalisations, and deaths reported across England over the course of the pandemic (Figure 1). In this study, we aim to characterise this heterogeneity in COVID-19 outcomes and how it evolved during the pandemic **(*1, 13, 14*)**. Similar studies on the impact of socioeconomic heterogeneities on SARS-CoV-2 infections have been conducted in France **(*15*)** and Germany **(*16*)** and on outcomes in the USA **(*17*)**.

**Figure 1:**
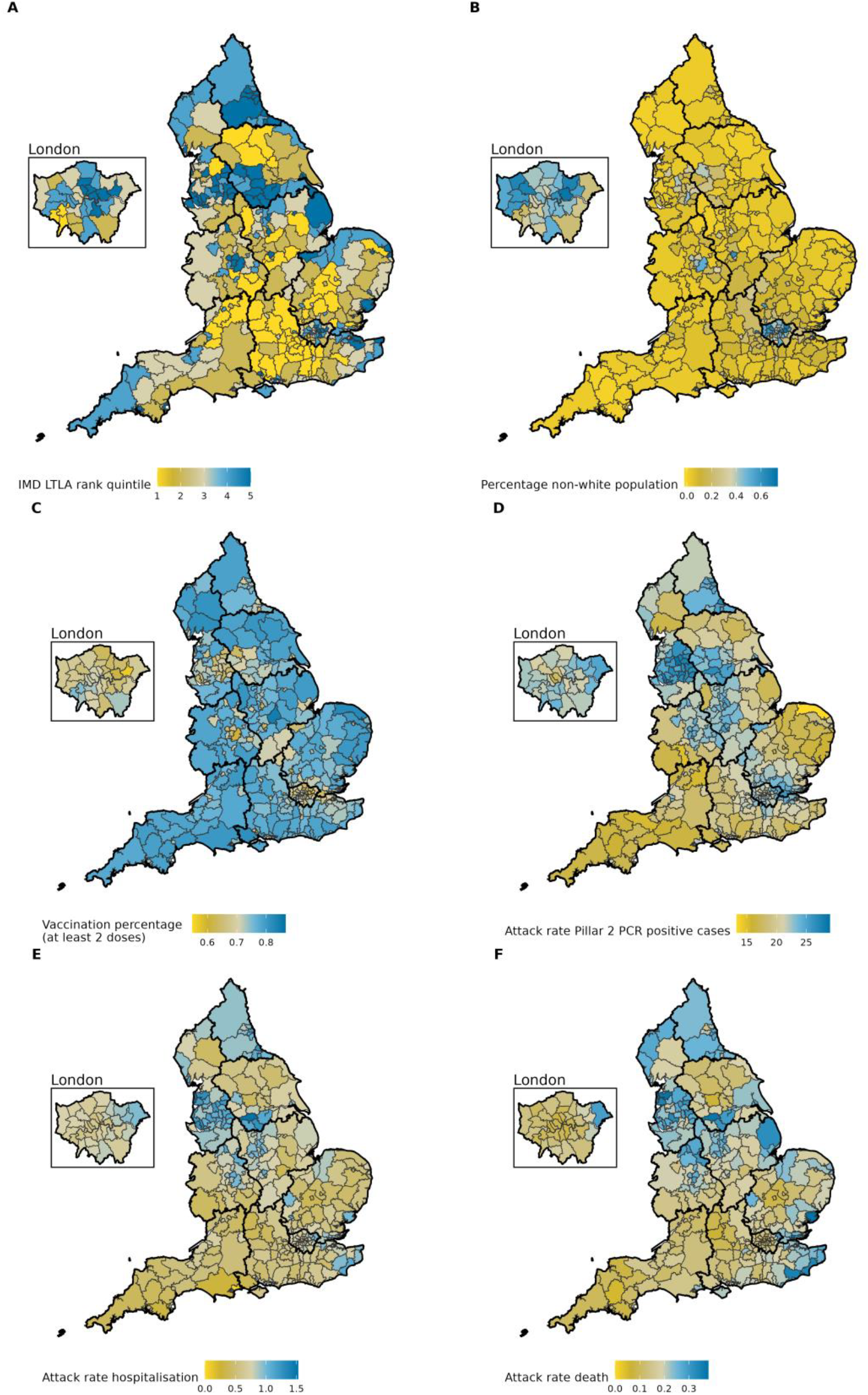
Maps of the population of England at the LTLA level between 10 May 2020 – 27 February 2022. (A) Index of Multiple Deprivation quintile by LTLA, (B) percentage of non-White population within the LTLA by LTLA, (C) percentage of individuals vaccinated with at least two doses for all ages, (D) percentage of positive Pillar 2 PCR confirmed first recorded cases by LTLA, (E) percentage of hospitalisation by LTLA, (F) percentage deaths by LTLA

SARS-CoV-2 testing in England **(*18*)** had four “Pillars”: Polymerase chain reaction (PCR) testing for health and care workers and individuals with clinical needs (Pillar 1), freely available PCR (and later antigen) testing for the general population (Pillar 2), serology (Pillar 3) and targeted surveillance (Pillar 4). Pillar 2 testing increased significantly from 10^th^ May 2020, and most testing was halted on 27^th^ February 2022 with the introduction of the ‘Living with COVID’ strategy **(*19*)** (see Methods and SI C.4).

We additionally investigate vaccine effectiveness (VE) in the post-vaccination period and the impact of vaccination on risk heterogeneity. The SARS-CoV-2 immunisation programme in England was one of the most rapid globally, with 89.9% of the adult population aged 20 or over receiving at least one dose and 86.6% receiving at least two doses by 27^th^ February 2022 **(*20*)**. Test-negative case-control studies (TNCCs) were the standard epidemiological tool for evaluating SARS-CoV-2 VE during the pandemic **(*21*–*23*)**, though TNCCs can potentially suffer from biases **(*24*)**. The main alternative to TNCCs used for VE estimation during the COVID-19 pandemic is large population observational cohort studies that use individual-level healthcare and surveillance data linked to vaccination status **(*25*–*28*)**.

Last, we investigate how risk heterogeneity was modified by the public health measures (“restrictions”) in force during the pandemic (see Methods and SI B.5) **(*1, 14*)**.

## Results

We present the breakdown of the England population (56,427,863 people in England [Census 2021], 56,417,353 are in our synthetic population, and 56,344,410 are included in the analysis) in Table 1, including the number of first recorded PCR-confirmed SARS-CoV-2 infections (12,310,485) and associated hospitalisations (439,083) or deaths (107,823) linked to that first recorded infection from the week beginning 10^th^ May 2020 (isoweek 18) to 27^th^ February 2022 (isoweek 8) confirmed by a Pillar 2 PCR positive test. These 95 weeks cover the full period for which population-wide testing was available.

**Table 1:**
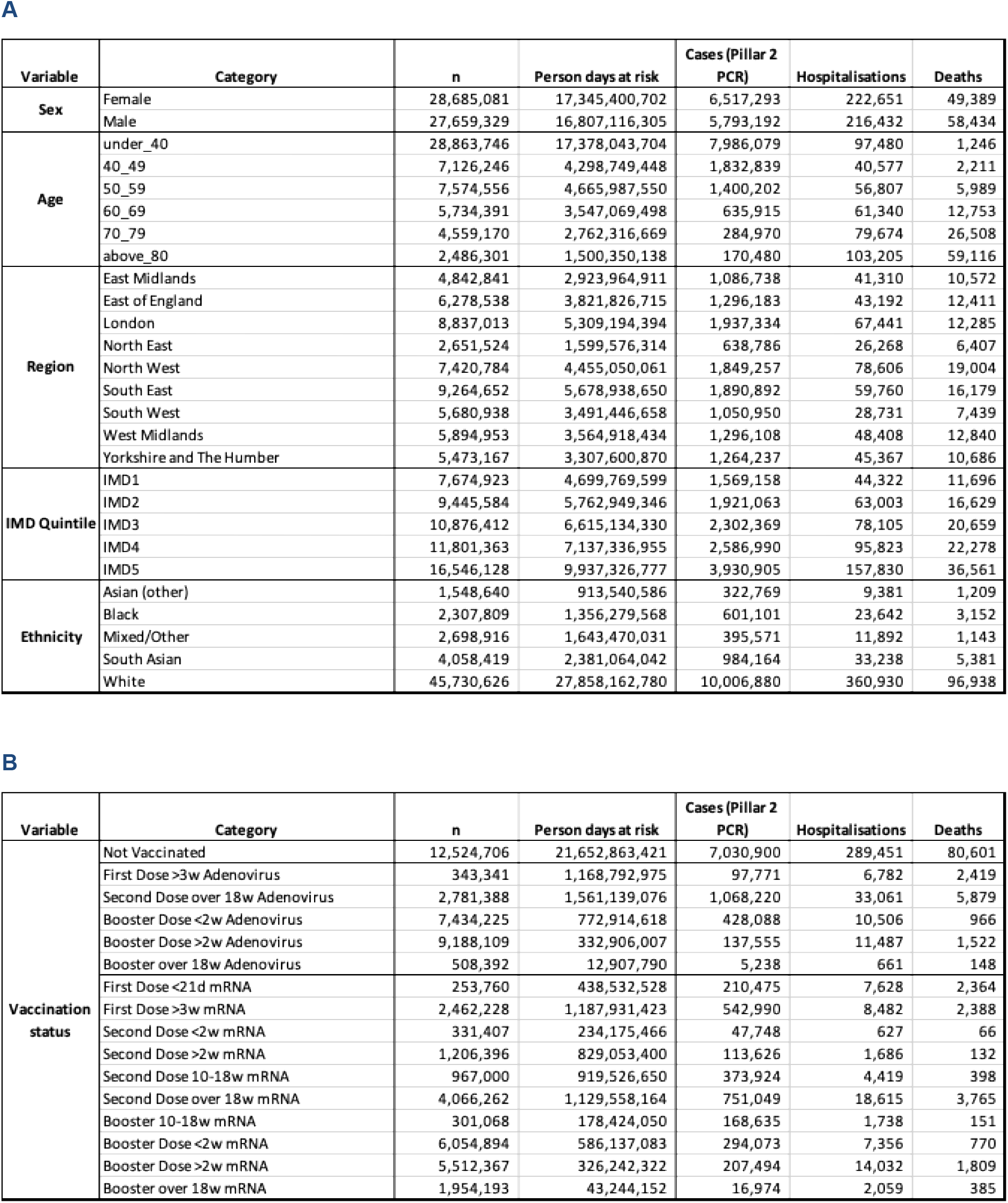
Population for England with numbers of first recorded Pillar 2 PCR confirmed SARS-CoV-2, hospitalisations and deaths in England between 10^th^ May 2020 and 27^th^ February 2022. (A) Population breakdown by sex, age, region, deprivation, and ethnicity. (B) Breakdown by vaccination status for categories with more than 200,000 doses at the end of the study period (n is the count of individuals in this category at the end of the study period, person days at risk and counts of Pillar 2 PCR positive cases, hospitalisations and deaths are over the entire study period). Vaccination status categories are by vaccination type (Adenovirus or mRNA), vaccine dose, and time since vaccination. The whole table for panel B is available in SI Figure D.9.

We examined a total of 360 model variants, which varied by the covariates and interactions included (see SM A.2 for covariate definitions considered): age, definition of deprivation, regional disaggregation, ethnicity disaggregation, and which interaction terms to include in the model. SM fig. S2-S4 lists the top 88 selected models for each outcome of interest. Restriction levels are based on individual policy measures, such as school closure or gathering restrictions, at each time point (SM section A, C.3 and fig. S7-S9). We found that less granular age categories had better 10-fold cross-validation performance (under 40s, 10-year age bands up to 70, single group for 80+). Similarly, coarse spatial disaggregation (by NHS region of England) was favoured, and deprivation was quantified by IMD quintiles. We disaggregated ethnicity into six groups as the results were very similar for the range of ethnicity categorisations examined. The preferred model included covariates for sex, age, NHS region, vaccine status, IMD quintile, ethnicity and restriction level.

We compared the estimates from the preferred model against the data and found a good qualitative fit examining marginal incidence rate ratios (IRRs) (SM fig. S1).

We investigated the impact of overdispersion for each outcome of interest using negative binomial regression models. For Pillar 2 PCR confirmed cases, we found a moderate level of overdispersion (*k*=0.33) during the wild-type period. We found that either very high values of *k* (indicating marginal overdispersion) or the Poisson model were preferred for all other periods. For hospitalisation and death outcomes, the Poisson model was always preferred (SM section A and table S1).

Figure 2 presents the modelled IRRs for deprivation and ethnicity categories from the preferred model for each endpoint and by period defined by variant dominance. IRRs for sex, age and region are presented in SM fig. S11.

**Figure 2:**
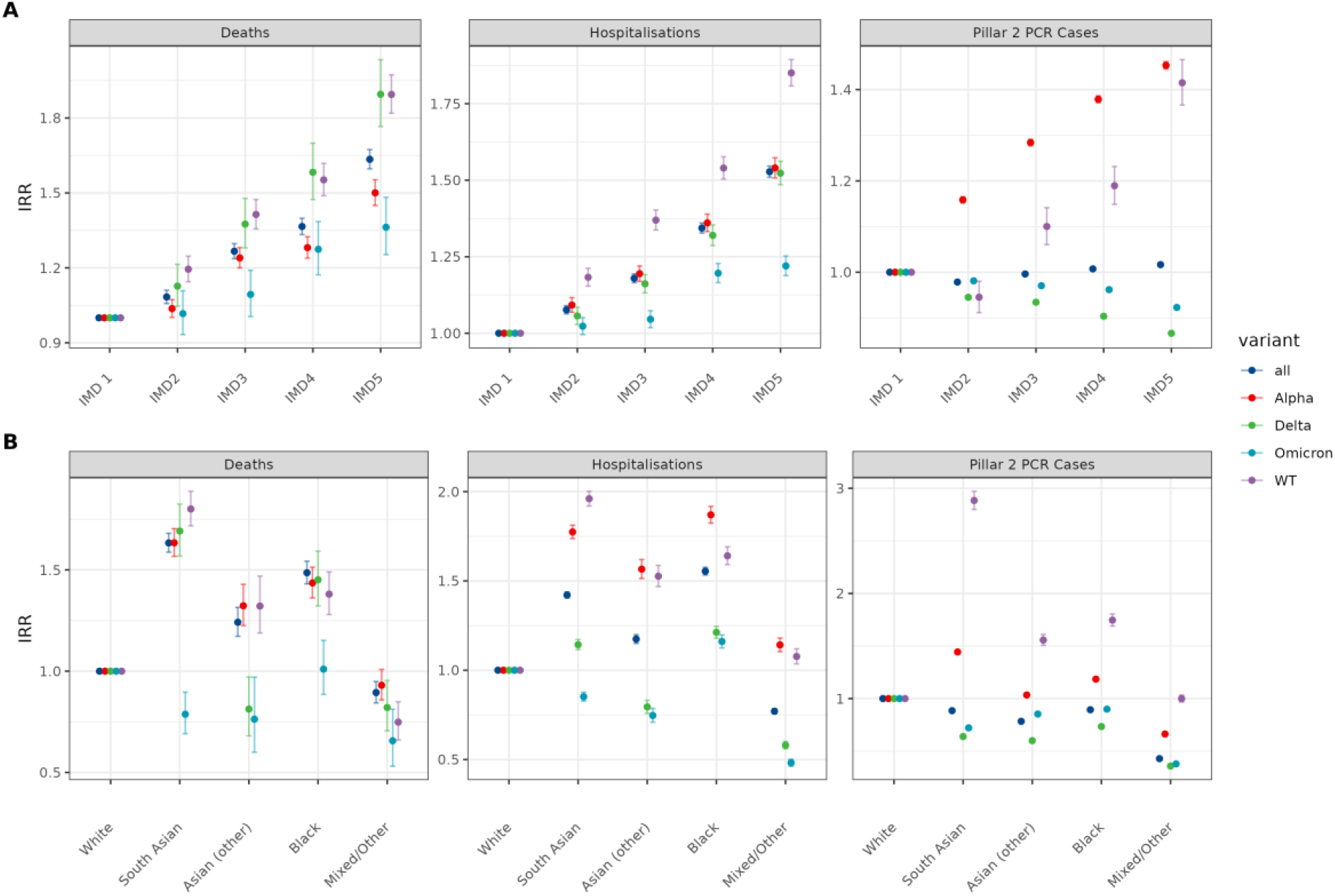
Estimated IRRs for (A) deprivation and (B) ethnicity covariates from preferred models. Deprivation was categorised by quintiles of the Index of Multiple Deprivation score defined at the LTLA level (results for sub-components are available in the SI), with the first (least deprived) quintile (IMD1) being the reference group (IRR=1). Results are shown for the whole pandemic (‘all’ - test dates between May 2020 and February 2022) and for the time intervals within that period where specific viral variants dominated (see SI for details). WT=Wild-Type (pre-December 2020).

When we consider the entire period (May 2020-February 2022), the highest IMD quintile, corresponding to the most deprived LTLAs, has an IRR (relative to the least deprived IMD quintile) of 1.64 (95% CI: 1.60-1.67) for death as the outcome, 1.53 (95% CI: 1.51-1.55) for hospitalisation, 1.02 (95% CI: 1.02-1.02) for a positive Pillar 2 PCR test. For the Pillar 2 positive test outcome, we observed significant differences between periods defined by variant dominance. The biggest differences were observed for the Alpha variant with an IRR (for the highest quintile relative to the lowest) for that endpoint of 1.45 (95% CI: 1.45-1.46).

For deaths, over the entire study period, South Asian ethnicity had an IRR 1.63 (95% CI: 1.59-1.68), Asian (other) ethnicity an IRR 1.24 (95% CI: 1.17-1.32), and Black ethnicity an IRR 1.49 (95% CI: 1.43-1.54), with White ethnicity as the reference. For hospitalisation, over the entire study period, South Asian ethnicity had an IRR 1.42 (95% CI: 1.40-1.44), Asian (other) ethnicity an IRR 1.17 (95% CI: 1.15-1.20), and Black ethnicity an IRR 1.56 (95% CI: 1.53-1.58). Using Pillar 2 PCR positive tests as the endpoint, over the entire study period, South Asian ethnicity had an IRR 0.89 (95% CI: 0.88-0.89), Asian (other) ethnicity an IRR 0.78 (95% CI: 0.78-0.79), and Black ethnicity an IRR 0.89 (95% CI: 0.89-0.90). The IRRs for Mixed/Other ethnicity are reported in SM table S4.

Ethnicity differences declined for the Pillar 2 PCR positive test endpoint throughout the pandemic. IRR for Pillar 2 PCR positive cases declines for all non-White ethnicities (relative to White) over time, with IRR_WT_ > IRR_Alpha_ > IRR_Delta_. During the Omicron period, non-White IRRs were broadly similar to the Delta period but typically slightly above IRR_Delta_. For death, we observed a declining IRR over time for South Asian ethnicity, with IRR_WT_, IRR_Alpha_, and IRR_Delta_ exhibiting overlapping confidence intervals but IRR_Omicron_ statistically significantly lower. For hospitalisation, we observed a declining IRR over time for South Asian ethnicity, with IRR_WT_, IRR_Alpha_, IRR_Delta_ and IRR_Omicron_ statistically significantly lower than in the previous period. Other Asian ethnicities had high IRRs (overlapping confidence intervals) for the WT and Alpha periods and significantly lower IRRs for Delta and Omicron. Black ethnicities exhibited high IRRs up to Omicron in the range of 1.38-1.45 for death and for the entire period for hospitalisation 1.16-1.87.

Results for deprivation were consistent for severe outcomes (hospitalisation and death) throughout the pandemic, with IRRs monotonically increasing as deprivation increased, but with IRRs during the Omicron period lower than other periods. This result also applied to a more granular representation of deprivation, using deciles (SM fig. S13) and for the different subcomponents of the IMD measure (SM fig. S20-S26). For Pillar 2 PCR positive cases, we observed an increasing IRR with increasing deprivation for the WT and Alpha periods but not for the Delta and Omicron periods. This resulted in the IRR for the whole period being close to 1. This result was consistent with IMD deciles and the subcomponents of the IMD index.

Figure 3 presents our VE estimates (SM section B.3 and table S3) for our preferred model. We obtain a VE (defined as 1-IRR) estimate for mRNA vaccines ranging from 86.8% (over 18 weeks after the second dose, 95% CI: 86.2-87.3) to 97.1% (less than two weeks after the booster dose, 95% CI: 96.6-97.6) for protection against death for individuals with at least two doses, 84.8% (over 18 weeks after second dose, 95% CI: 84.5-85.0) to 93.3% (2-10 weeks after booster dose, 95% CI: 93.1-93.5) against hospitalisation and 30.4% (over 18 weeks after booster dose, 95% CI: 29.3-31.5) to 84.5% (2-10 weeks after second dose, 95% CI: 84.4-84.6) against Pillar 2 PCR positive confirmed infections for the full study period.

**Figure 3:**
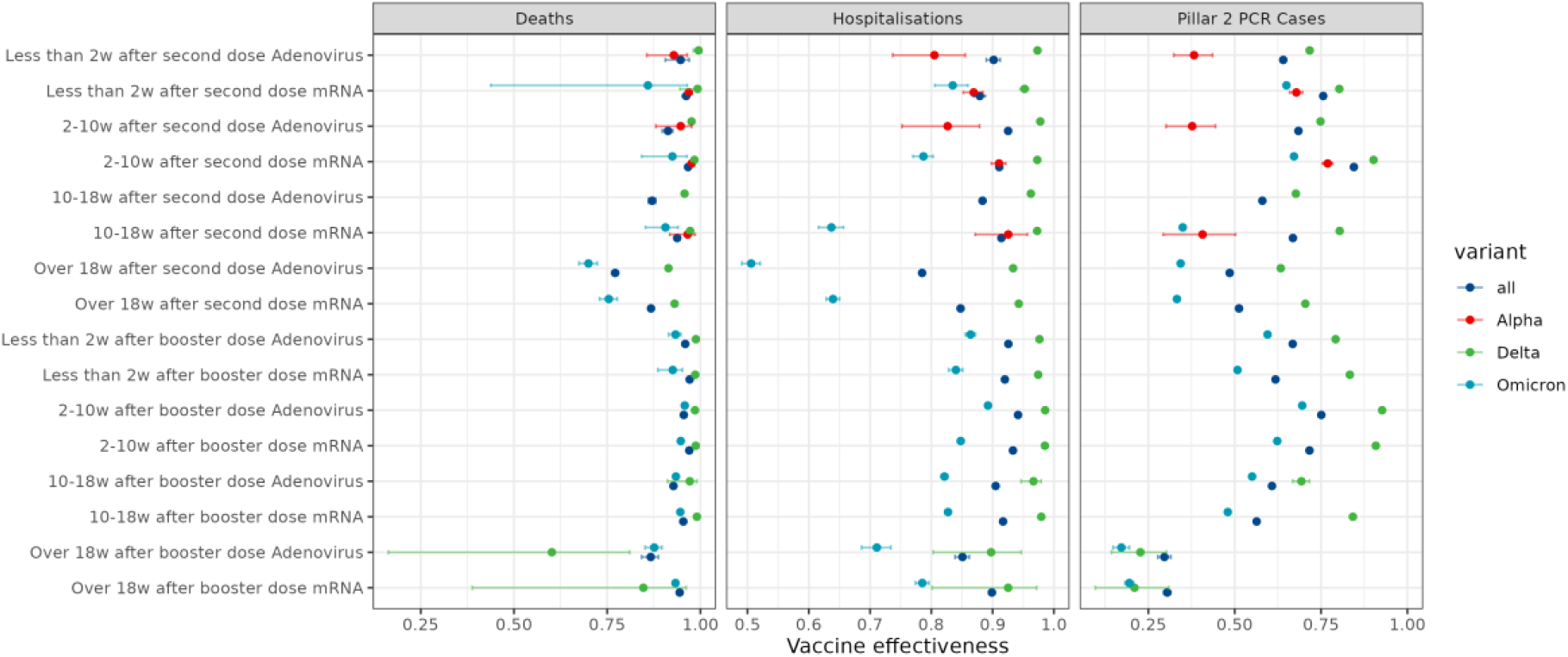
Vaccine effectiveness (VE=1-RR) for different vaccination statuses with not vaccinated as the reference group (IRR=1 / VE=0). Results are shown for the whole pandemic (‘all’ - test dates between May 2020 and February 2022) and for the time intervals within that period where specific viral variants dominated (see SI for details). Results for the Second Dose and Booster Dose categories are displayed for mRNA and Adenovirus vaccines (all other results are in SI Table D.3 and Figure D.11). WT=Wild-Type (pre-December 2020).

For adenovirus-based vaccines, we estimated VE for individuals with at least two doses, ranging from 77.2% (over 18 weeks after second dose, 95% CI: 76.2-78) to 95.9% (less than two weeks after booster dose, 95% CI: 95.3-96.5) for protection against death, 78.5% (over 18 weeks after second dose, 95% CI: 78.2-78.8) to 94.1% (2-10 weeks after booster dose, 95% CI: 94.0-94.3) against hospitalisation, and 29.6% (over 18 weeks after booster dose, 95% CI: 27.6-31.5) to 68.5% (2-10 weeks after second dose, 95% CI: 68.3-68.6) against Pillar 2 PCR-positive confirmed infections for the full study period.

We found that VE was consistently lower for the Omicron period. There was little to no difference in VE estimates for severe outcomes for the Alpha and Delta time periods.

We present the estimated VE results for all variants, vaccine status, and vaccine types in SM table S3 with their associated 95% CIs. The confidence intervals are narrow due to the large number of events observed (Table 1B); confidence intervals are wider for the variant period-specific estimates. A comparison with previous vaccine effectiveness estimates is included in SM section D.2.

Figure 4 shows the estimated VE results obtained from an extension of the preferred model, which included an additional interaction term between vaccine status and IMD quintile. Panel (A) presents results for the period when Delta was the dominant variant, while panel (B) covers Omicron. For deaths and hospitalisations, we did not observe significant differences in VE between IMD quintiles, except for the period over 18 weeks post-second dose during the Delta phase for deaths, where there was a tendency for the least deprived areas (IMD 1 and 2) to have lower VE than the more deprived areas (IMD 4 and 5).

**Figure 4:**
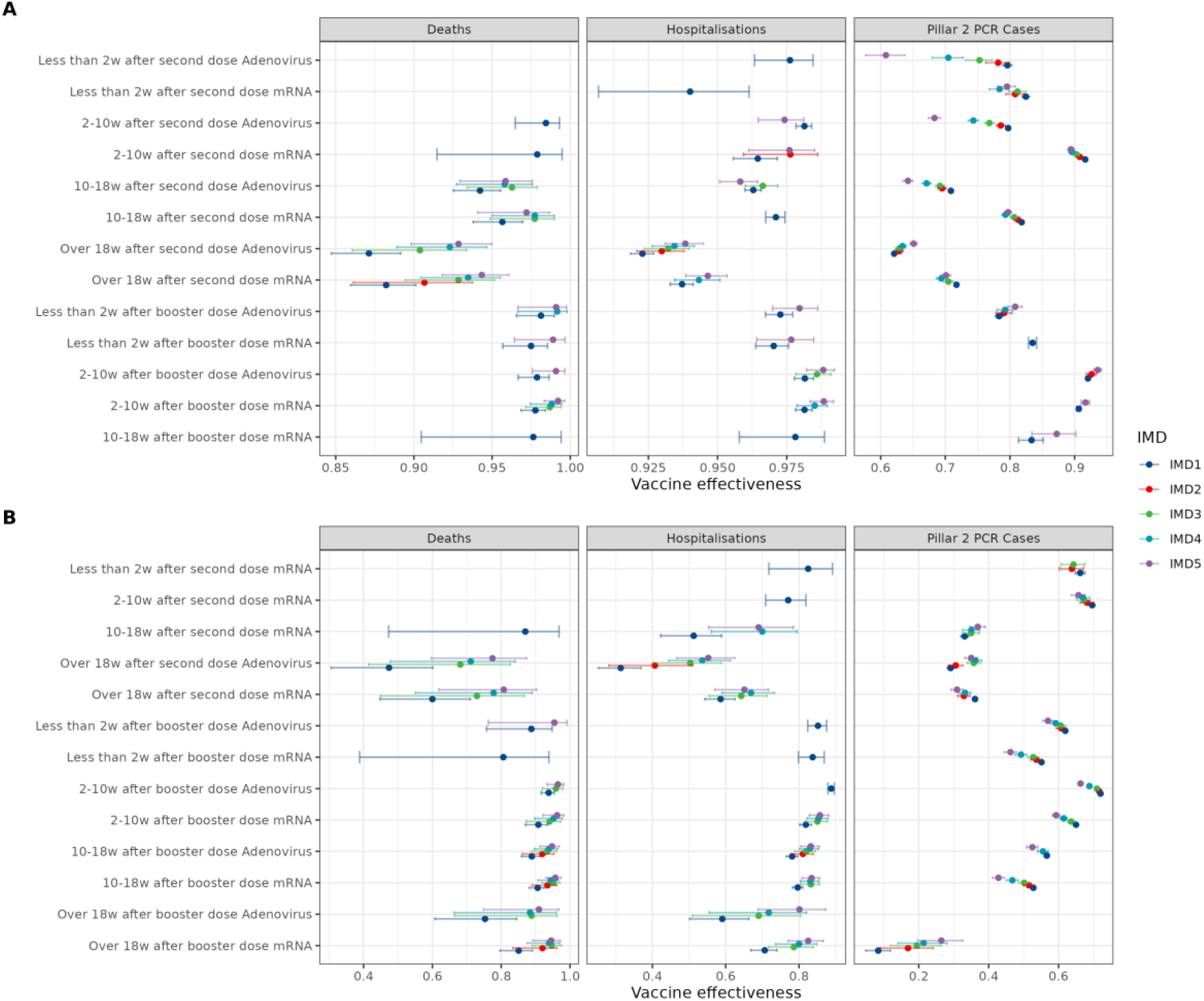
Vaccine effectiveness (VE=1-RR) for the preferred model with an additional interaction term for vaccine status and IMD, with not vaccinated as the reference group (IRR=1 / VE=0). Results are shown for the Delta period in panel (A) and Omicron period in panel (B), and results for the Second Dose and Booster Dose categories are displayed for mRNA and Adenovirus.

For PCR-confirmed infections, we found different results for the Delta and Omicron periods. During Delta, we typically observed the ordering of IMD quintiles was higher VE for less deprived areas than more deprived areas, particularly less than 18 weeks since the second dose. VE estimates cluster together for over 18 weeks since the second and booster doses. For Omicron, we observed little variability in VE by IMD for second and booster doses, but VE estimates were higher for less deprived areas. The exception is for over 18 weeks since the booster dose, for which we observed higher VE for more deprived areas. Results for IRRs of all covariates for the model with an interaction term between vaccine status and IMD quintile are presented in SM fig. S17 and for vaccine status and ethnicity in SM fig. S18.

Figure 5 shows the estimated IRRs for the restriction level covariate for our preferred model. The IRRs for restriction levels were robust to regional resolution, up to and including the LTLA level (SM fig. S15 and S16). We estimated the IRR for restriction level over the full study period of 1.14 (95% CI: 1.12-1.15) for deaths, 1.15 (95% CI: 1.14-1.16) for hospitalisation, and 1.17 (95% CI: 1.17-1.17) for Pillar 2 PCR positive cases. These estimates indicate that periods of high levels of public health restrictions were associated with higher levels of risk than periods where restriction levels were lower. The estimated IRR for restriction level for the WT and Alpha periods was lower for deaths and hospitalisation than for the full-time period, which includes longer time intervals with no restrictions. However, for Pillar 2 PCR-confirmed infections, we found that the IRR for restriction level was higher during the WT period and lower for the Alpha period than for the full-time period (Figure 5). The IRRs for the Delta and Omicron periods are one, as no substantial restrictions were in place over those periods.

**Figure 5:**
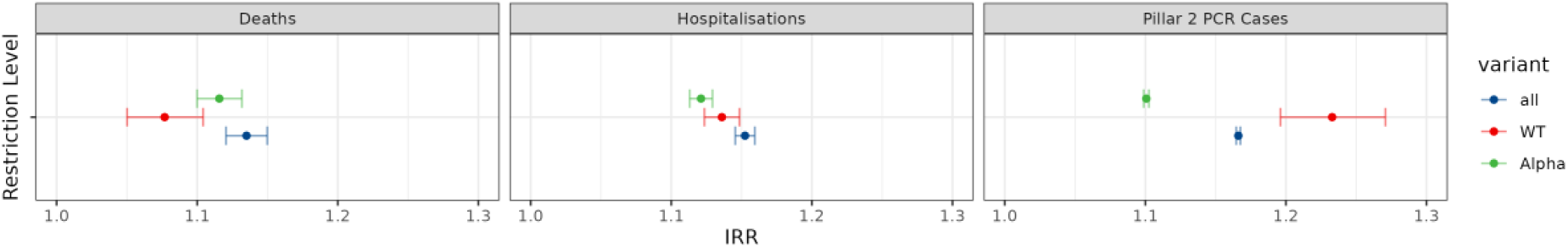
Estimated IRRs for restriction levels. Results are shown for the whole pandemic (‘all’ - test dates between May 2020 and February 2022) and for the time intervals within that period where specific viral variants dominated (see SI for details). Restriction level is a numeric covariate in the model.

## Discussion

We considered the impact of sex, age, region, deprivation, level of restrictions, and ethnicity on the relative risk of a SARS-CoV-2 Pillar 2 PCR-positive first recorded infection, hospitalisation, or death. Our proposed model was selected to provide a good fit across all three outcomes of interest. Our results extend previous work on the impact of deprivation **(*5*)** and evidence risk heterogeneity by ethnicity in England **(*4, 6, 8*)** and globally **(*7, 17*)**.

We find that modelling overdispersion for Pillar 2 PCR positive cases was necessary for the wild-type period, consistent with Morrissey et al. **(*13*)**, and the value for the overdispersion we find aligns with other estimates outside China **(*29, 30*)**. We did not find overdispersion to be present for severe outcomes, and the Poisson model provides the best fit to the data. Even though Pillar 2 PCR tests were available widely, this indicates that some groups, defined by higher levels of deprivation or non-White ethnicity, may have accessed testing less, leading to the observed overdispersion. This was less likely to be the case for more severe outcomes as testing would be more likely conducted due to the more severe nature of the infection.

The differences in IRRs we observe by outcome (PCR-confirmed infection, hospitalisation, or death) indicate a difference between severe outcomes and infections. For severe outcomes, we observed a significant risk heterogeneity by age, with older age groups at higher risks than younger age groups at an exponential rate (SM fig. S11(B)) consistent with the variation of the infection fatality ratio by age in the pre-vaccination era **(*31*)**. Increased risk was also positively associated with higher levels of deprivation (Figure 3A), and the difference between the most deprived quintile of LTLAs and the least deprived quintile was significant for all periods considered. Similarly, we found that non-White ethnicities were at increased risk.

For Pillar 2 PCR-positive cases, the picture is more nuanced. We observed that the differences in risk between age groups were of a similar magnitude as the large differences we observed for severe outcomes. These differences are consistent with past studies and may reflect age-specific self-protective behaviours **(*32*)** and/or variation in contact rates (SM fig. S6). For deprivation and ethnicity, risk heterogeneity was similar in trends as for more severe outcomes, but the magnitude of differences was less. The results for Pillar 2 PCR-positive cases were consistent with other studies **(*32, 33*)**.

For severe outcomes, we observed less variation in deprivation or ethnicity related risk differences across the course of the pandemic, pointing to existing health inequalities driving the strong association between deprivation, ethnicity, and risk of severe outcomes. This held consistently for all sub-components of the index of multiple deprivation, except for the Living Environment component, and particularly strongly for the Income, Employment, Health, and Education sub-components (SM fig. S20-S26). This observation is consistent with known socio-economic and ethnic disparities in the UK for type-2 diabetes (*34*), which is a known risk factor for severe SARS-CoV-2 infection outcomes. Interestingly, we observed a high IRR for South Asian ethnicity, for both severe outcomes and infections, which reduced over time (Figure 3.B), perhaps suggesting the metabolic component impacting outcomes being reduced for omicron and delta as individuals of South Asian ethnicity are at higher risk of metabolic disease (*35*).

We assessed vaccine effectiveness, and our results broadly align with previously published VE estimates for England by UKHSA(*21*) for similar periods and VE definitions. We find vaccine effectiveness for the mRNA vaccines is consistent with UKHSA estimates of 88.0% (95% CI: 85.3-90.1) for two doses of BNT162b2 for infections for persons with the Delta variant. Similarly, we find vaccine effectiveness for the adenovirus vaccines is in line with UKHSA estimates of 67.0% (95% CI: 61.3-71.8) for persons with two doses of the ChAdOx1 nCoV-19 vaccine. For the Omicron variant, estimates vary more widely(*36*) with VE against hospitalisation, ranging from 50.5% to 89.2%, broadly consistent with 67.8% to 92.4% from the literature. Similarly, VE against Pillar 2 PCR confirmed infection ranged from 17% to 69.5% compared to 11.7% to 75.3% in the literature. We note that we only consider the first recorded infections for the Omicron period, which may bias our VE estimates to be higher, which is offset by our longer study period.

We considered VE stratified by deprivation (Figure 4). For Pillar 2 infections, observed differences in VE may be explained by differences in the propensity to seek testing which was associated with both deprivation level and vaccination status. Later in the pandemic, the propensity to seek testing was substantially lower in the most deprived areas than the least deprived (*37*). Higher VE for the least deprived areas might also be due to higher levels of infection-induced immunity in more deprived areas; if mild infections were less likely to be detected in more deprived areas, the unvaccinated group would have higher immunity levels, resulting in lower VE estimates for the most deprived IMD quintile.

The policy implications of our study are that both protective and support measures need to be structured with ethnicity and deprivation in mind, particularly in the early stages of a pandemic. Deprivation had a particularly significant impact during the Alpha variant era for PCR-confirmed infections. For ethnicity, we observed a reduction in IRRs as the pandemic progressed (Figure 3). This suggests that more deprived areas were less able to isolate effectively, potentially due to work, during heightened public health restrictions. Ethnic minorities were more likely to work in occupations where workplace attendance continued even during lockdowns and also had less access to savings (*38*), potentially exposing them to a higher risk of community transmission. This is particularly true of, Bangladeshi and Pakistani men working in the hospitality, leisure and transport sectors. South Asian and African-Black men are also overrepresented in health and social care roles in England, potentially leading to higher levels of infection risk during the pandemic (*38*).

Considering the impact of the public health restriction level, we found IRRs>1 for all outcomes in the wild-type and Alpha periods. Restrictions were highest during periods of high hospitalisation and death incidence (SM fig. S8). The higher IRR for deaths during Alpha, relative to Wildtype, was consistent with the hospitalisation fatality ratio (HFR) being higher for Alpha than Wildtype (*39*). The IRR for deaths over the entire study period was comparable to the IRR for deaths during Alpha, suggesting that vaccination during Delta and Omicron, both periods of low to no restrictions, contributed to lower risk relative to Wildtype and Alpha. We find an IRR greater than one for hospitalisations but higher IRRs during Wildtype than Alpha. Similarly to deaths, we find the highest IRRs for the entire study period, pointing to the impact of vaccination in the second half of the study period. For Pillar 2 PCR cases, we observe an IRR greater than one, with IRRs highest during Wildtype. The IRRs for the full study period are lower than for Wildtype, as we observed significant infections in both the unvaccinated and vaccinated groups during Omicron, a period with no restrictions. We note that this IRR for the entire study period would be lower if we had taken Lateral Flow Tests (LFTs) into account.

This highlights the impact of the different timing of restrictions between regions, as we already account for week and region effects in the model. We note that other results were not sensitive to whether restrictions were included as a predictor in the model, and the IRRs for restrictions were robust to different resolutions of the regional covariate.

A limitation is that we only considered first recorded infections in this study and did attempt to investigate reinfections. However, we note that up to November 2021, with the arrival of the Omicron variant of SARS-CoV-2, 93% of all recorded infections were first infections. In addition, we only quantify incidence among those seeking tests, not underlying infection incidence. This may impact our results if the propensity to seek a test varied by age, deprivation or ethnicity, especially if underlying true infection incidence varied substantially by those variables. Hence for the Pillar-2 test outcome, risk and VE differences between population groups/strata may reflect true exposure differences, differences in propensity to seek a test, or both. Such considerations do not affect the hospitalisation and death endpoints, as testing was universal for anyone entering hospital for the large majority of the study period. For Omicron, we only consider PCR test results and do not include Lateral Flow Tests (LFTs).

Future research priorities include a multi-outcome survival model to incorporate infections and severe outcomes in the same model. Integrating serology and infection prevalence data is essential in addressing questions such as the impact of pre-existing comorbidities on the risk heterogeneity by ethnicity or socio-economic group.

## Material & Methods

### Data sources

The UK Health Security Agency (UKHSA) maintains a single unified database of all SARS-CoV-2 positive test results in the country, categorised by Pillar. The resulting database contains a hashed version of a unique identifier (NHS number), the age, sex, ethnic group, home LTLA, symptom status, testing Pillar, and date of specimen for each positive case.

Second, linked to the case database by (hashed) National Health Service (NHS) number, UKHSA maintains a database of all deaths within 28 days of a positive PCR test for SARS-CoV-2. This database records the date of death, the date of hospital admission, and how the death was identified (e.g., via hospital reporting, death registration, or both).

Third, two NHS data sources on hospitalisation were also linked by NHS number to the case database: the Secondary Uses Service (SUS) data on hospital episodes and the Emergency Care Data Set (ECDS) on attendance at Accident and Emergency departments. An individual in these datasets was classified as hospitalised with COVID-19 if they had a positive PCR test between 14 days before admission and the day before discharge, and they were classified as an inpatient.

Lastly, England maintains a national vaccination register, the National Immunisation Management System (NIMS) (*20*). Every SARS-CoV-2 vaccine dose given in England is recorded at the time of vaccine administration. The anonymised version of the dataset used here contained a unique identifier (hashed NHS number), age, sex, ethnic group, LTLA and administration date.

We extracted data from the versions of these databases with data up to 25^th^ July 2023. Cases with a valid NHS number were then linked to the NIMS vaccination dataset. Where a match was found, the case was classified as having received a vaccine (either before or after testing positive). Cases with no matching immunisation record were classified as having not been vaccinated. In our analysis, we used the vaccination status of an individual at the time of testing. The vaccination status includes the type of vaccine, number of doses received and time since the last vaccination.

PCR testing (Pillars 1 and 2) was initially focused on key workers in the NHS, social care and other sectors due to limited testing capacity. From May 2020, testing was widened to individuals with symptoms in the general public and regular asymptomatic testing of individuals working in high-risk settings, such as care homes, was introduced **(*40*)**. Individuals with a positive SARS-CoV-2 test were initially advised to self-isolate for 14 days, with subsequent rules being more complex and dependent on testing (fig. S9). Limited support was available to individuals who had to self-isolate, provided they met several criteria, such as not being able to work at home and being in receipt of at least one state benefit **(*41*)**.

Population denominator data was used to calculate the number of people who have neither received a vaccine nor been diagnosed with SARS-CoV-2 infection. We used the 2021 Census population estimates generated by the ONS (*42*), stratified by age, sex and LTLA (*43*). We made the simplifying assumption that in the absence of COVID-19, the size of the English population and its age distribution would have been at a steady state for our analysis period consistent with the Census 2021 estimates. Individuals with missing information on sex, age, or residence location were not included in the analysis.

We use the Index of Multiple Deprivation (IMD) dataset released in 2019 (*12*) by the UK Ministry of Housing, Local Communities, and Local Government, aggregated at the LTLA level and its subcomponents (SM section B.4).

The public health and social restrictions level was computed from ONS LTLA data up to December 2020 (*44*) and the Oxford COVID-19 Government Response Tracker (*1*) (SM section A and C.3). Several non-pharmaceutical interventions (NPIs) were used across the course of the pandemic, including containment, school closure and stay at home orders. The “roadmap out of lockdown” policy in spring 2021 had three intermediate steps between national lockdown and no restrictions. We map the restrictions over time onto a 5-level scale based on the individual policy measures in place at each time point (SM section C.3).

### Statistical Analysis

Survival models were used to estimate Incidence Rate Ratios (IRR), an estimate of the increased risk of a particular outcome for a specific population group relative to a reference group. We used Poisson regression using an offset to adjust for person-days at risk to generate IRRs for the different strata of interest, proving a good approximation to continuous time survival models (SM section A)(*45*). Poisson regressions are more general than Cox proportional hazard survival models, allowing for non-proportional hazards. We investigated using negative binomial regressions to account for overdispersion. We parametrically captured variation by systematically examining evidence for interactions between covariates. We conducted model selection to identify the model which performed best across all three outcomes of interest (infection, hospitalisation, and deaths) by considering the average Akaike’s information criterion (AIC) and coefficient of determination (R^2^) (*46, 47*) across the three respective models.

We included terms for sex, ethnic group, vaccination status and IMD in all models. We used a categorical IMD variable, ranking LTLAs by average IMD score and then binning them into quintiles (1 being least deprived, five being most). The four other covariates included in all models were geographic region (either upper tier local authority or region), age (in 10-year bands with the last band being 85+ or 10-year bands with all individuals below 40 grouped and a max-age of 80+), restriction levels (used as a parametric covariate as restriction levels changed several times, creating non-continuous time intervals) and epidemiological week. We examined all pairwise interactions between these three variables. The combined restriction covariate was also included in the preferred model.

All survival analyses used the test specimen date as the outcome event date. Three SARS-CoV-2-related events were defined: death within 28 days of any positive PCR test, hospitalisation where the individual hospitalised tested PCR-positive between 14 days before admission and the day before discharge and was classed as an inpatient, and any PCR-confirmed infection within Pillar 2. We only consider the first recorded SARS-CoV-2 infection for all three events and associated hospitalisations and deaths.

We fitted the model for all three outcomes of interest. Individuals were treated as censored after a positive SARS-CoV-2 PCR test to exclude post-primary infections and avoid bias since the UK vaccination programme excluded people from being vaccinated within 28 days of a positive test.

Since the UKHSA/NHS and ONS data used different ethnicity categories, we aggregated ethnicities into six groups that could be identified consistently across all datasets: White, South Asian, Asian (other), Black, and Mixed/Other to be consistent across two datasets (SM section B.1). Details are provided in Supplementary Materials (SM section B.1). The computation of age for each individual is outlined in the SM section B.2.

We estimated VE accounting for the type of vaccine (mRNA or adenovirus-based), time since vaccination, and outcome of interest (death, hospitalisation, Pillar 2 PCR positive case). Details are provided in SM section B.3. By comparing currently vaccinated individuals against people who would be vaccinated in the future, we controlled for potential differences in behaviour and exposure risk between those who are never vaccinated and those who eventually are. Censoring after the first recorded positive test partially mitigated estimates from being affected by the accumulation of naturally acquired immunity in the population.

To investigate how estimates varied over time, for example, in periods dominated by particular variants, we restricted model fitting to data from the specific period of interest (censoring individuals with positive tests before that period).

We used negative binomial regressions as a sensitivity analysis to explore over-dispersion in the count data. In all figures, we display only results with a p-value of 0.05 or less.

Interactions between VE and deprivation were examined by running the preferred model with an additional interaction term for vaccine status and IMD. Similarly, we investigated associations between VE and any ethnicity by running the preferred model with an interaction term for vaccine status and ethnicity.

All analyses were undertaken in R version 4.3.1 using the H2O.ai machine learning package version 3.42.0.2 (*48*), which offers high-performance parallelised algorithms for fitting general linear models to large datasets. A 32-core Intel-based server with 128GB RAM was used to conduct the analyses.

## Data Availability

While all data used in this analysis were anonymised, the individual-level nature of the data used risks individuals being identified or being able to self-identify if it is released publicly. Requests for access to the underlying source data should be directed to UKHSA.

## Data access

While all data used in this analysis were anonymised, the individual-level nature of the data used risks individuals being identified or being able to self-identify if it is released publicly. Requests for access to the underlying source data should be directed to UKHSA. The access to protected data request is available at https://www.gov.uk/government/publications/accessing-ukhsa-protected-data/accessing-ukhsa-protected-data. All other data needed to evaluate the conclusions of the paper are present in the paper and/or the Supplementary Materials.

## Ethical approval

Surveillance of COVID-19 testing and vaccination is undertaken under Regulation 3 of The Health Service (Control of Patient Information) Regulations 2002 to collect confidential patient information (www.legislation.gov.uk/uksi/2002/1438/regulation/3/made) under Sections 3(i) (a) to (c), 3(i)(d) (i) and (ii) and 3(3). Data were shared with the investigators as part of the UK’s emergency response to the COVID-19 pandemic via the SPI-M subcommittee of the UK Scientific Advisory Group for Emergencies (SAGE). Ethics permission was sought for the study via Imperial College London’s standard ethical review processes, and the study was approved by the College’s Research Governance and Integrity Team (ICREC reference: 21IC6945).

## Role of the funding source

The funders of the study had no role in the study design, data collection, data analysis, data interpretation, or writing of the report.

References from SI (need to be at the end of the main text references; delete these lines once unlinked for resubmission): (*1, 12, 14, 21, 42, 45, 48, 49*)

## Declarations

### Code availability

https://github.com/cm401/covid19_risk_heterogeneity_england (https://doi.org/10.5281/zenodo.15202350)

### Authors’ Contributions

Conceptualisation: NMF, CM, SB

Data curation & validation: CM, WH, PPG, NMF

Methodology: CM, TR, WH, PPG, SB, NMF

Investigation: CM

Visualisation: CM

Supervision: NMF, SB

Writing – original draft: CM

Writing – review & editing: CM, TR, WH, PNPG, SB, NMF

All authors discussed, edited, and approved the final version of the manuscript. All authors had final responsibility for the decision to submit the manuscript for publication.

