## Supplementary material for "Socioeconomic and temporal heterogeneity in SARS-CoV-2 exposure and disease in England from May 2020 to February 2023": SARS_CoV_2_risk_heterogeneity_England_SI_medrxiv.pdf

Christian Morgenstern *et al*

March 2025

In section A of the SI, we provide more details on the survival model used and how it is estimated. Section B provides information on the time periods used, how we define sub-periods in the analysis, and how we consider restrictions. Section C provides additional figures for the results presented in the main text. Section D analyses the sub-components of the Index of Multiple Deprivation.

### A Survival model

A Poisson representation of the survival data likelihood is set out in section 3 of Laird et al [1]. Let $\log \hat{\rho}$ be the logarithm of the estimated person-year rate, which we define by Equation (1) as the difference of the logarithm of the number of events $\log e$ and the logarithm of the number of person-years at risk $\log y$.

Equation (2) represents the regression model where $x_i$ is a categorical variable representing a group membership (for example for sex this would be male and female). $r_i$ is a numeric variable encoding the level of restrictions present (see the section on NPIs C.3).

$$\log \hat{\rho} = \log e - \log y \tag{1}$$

$$\log \hat{\rho} = \beta_0 + \sum_{i \in strata} \beta_i \cdot x_i + \beta_k \cdot r_i \tag{2}$$

We can rewrite Equation (2) as

$$\log e = \log y + \beta_0 + \sum_{i \in strata} \beta_i \cdot x_i + \beta_k \cdot r_i \tag{3}$$

which allows us to consider this as a general linear model with a log link and the number of person-years at risk as an offset.

All analysis was undertaken in R version 4.3.1 using the H2O.ai version 3.42.0.2 machine learning library [2], since the latter offers high-performance parallelised algorithms for fitting general linear models to very large datasets. A 32 core Intel-based server with 128GB RAM was used to conduct the analyses.

We have a choice to consider that the events under consider ration could be either Poisson or Negative Binomial distributed. If $e \sim Poisson$ the model is fitted by maximizing the following penalised likelihood for the model in Equation (4):

$$\hat{y} = e^{x^\top \beta + \beta_0} \tag{4}$$

$$\max_{\beta_0, \beta} \frac{1}{N} \sum_{j=1}^{N} \Big( y_j \big( x_j^\top \beta + \beta_0 \big) - e^{x_j^\top \beta + \beta_0} \Big) - \lambda \Big( \alpha \left\| \beta \right\|_1 + \frac{1}{2} (1 - \alpha) \left\| \beta \right\|_2^2 \Big) \tag{5}$$

with the corresponding deviance equal to $D = -2 \sum_{j=1}^{N} \big( y_j \log(y_j / \hat{y_j}) - (y_j - \hat{y_j}) \big)$. We note that Equation (4) is the same as Equation (2) as $\sum_{i \in strata} \beta_i \cdot x_i = x^\top \beta$.

If we have overdispersion of events it is more appropriate to consider $e \sim Negative\, Binomial$, which is a generalisation of the Poisson regression loosening the assumption that the mean and variance have to be equal. Consider $\sigma^2 = \mu + \theta \mu^2$, where $\theta$ is the overdispersion parameter.

In the negative binomial case we have the probability for observation $i$ given by

$$Pr(Y = y_i | \mu_i, \theta) = \frac{\Gamma(y_i + \theta^{-1})}{\Gamma(\theta^{-1}) \Gamma(y_i + 1)} \left( \frac{1}{1 + \theta \mu_i} \right)^{\theta^{-1}} \left( \frac{\theta \mu_i}{1 + \theta \mu_i} \right)^{y_i} \tag{6}$$

where $\Gamma$ is the gamma function and $\mu_i = e^{x^\top \beta + \beta_0}$ as we consider a log link function in our use case.

The negative log-likelihood is given in Equation (7)

$$L\big(y_i, \mu_i\big) = \max_{\beta, \beta_0} \left[ \frac{-1}{N} \sum_{i=1}^{N} \left\{ \left( \sum_{j=0}^{y_i - 1} \log(j + \theta^{-1}) \right) - \log(\Gamma(y_i + 1)) - (y_i + \theta^{-1}) \log(1 + \theta \mu_i) + y_i \log(\mu_i) + y_i \log(\theta) \right\} \right] \tag{7}$$

And we get the penalised log-likihood as $L(y_i, \mu_i) + \lambda\big(\alpha \left\|\beta\right\|_1 + \frac{1}{2}(1-\alpha)\left\|\beta\right\|_2\big)$ corresponding to the deviance given by $D = 2\sum_{i=1}^{N}\Bigg\{y_i \text{log}\big(\frac{y_i}{\mu_i}\big) - (y_i + \theta^{-1})\text{log}\frac{(1+\theta y_i)}{(1+\theta\mu_i)}\Bigg\}$. We note that this is for a *given* value of $\theta$, that is we can't jointly infer $\beta$ and $\theta$.

In order to assess which model is more appropriate for our data, either Poisson or Negative Binomial, we estimate the Poisson model and Negative Binomial model for a range of $k$:
$k = \theta^{-1} = \{1e-04, 0.001, 0.01, 0.2, \frac{1}{3}, 1, 2, 3, 10, 100, 1000\}$ and we note that the Poisson case is equivalent to $k = \infty$.

| **Time period** | **Pillar 2 PCR cases** | **Hospitalisations** | **Deaths** |
|---|---|---|---|
| May 2020 - Feb 2024 | 1000 | Poisson | Poisson |
| WT | $\frac{1}{3}$ | Poisson | Poisson |
| Alpha | 100 | Poisson | Poisson |
| Delta | 1000 | Poisson | Poisson |
| Omicron | 1000 | Poisson | Poisson |

Table S1: We estimated the main model of interest, as described in the main text, for both the Poisson model and the Negative Binomial model for a range of values of $k$. The AIC of the out-of-sample prediction for 10-fold cross validated models was used for the selection of $k$.

Pillar 2 PCR positive cases (first infections) exhibit overdispersion and we use $k = \frac{1}{3}$, corresponding to $\theta = 3$ for the wild-type period in the results which we present. For associated hospitalisations and deaths we find that the Poisson model provides the best fit, consistent with our prior view that we should observe little or no overdispersion for either hospitalisations or deaths. For Pillar 2 PCR positive cases for all other periods, as well as the overall time-period, we use the Poisson model as estimates of $k$ are large and AICs similar to the Poisson model.

#### A.1 Model validation

In Figure S1 we present the marginal IRRs computed directly from the input data and compare these to the IRRs computed from our best model as a visual indication of how well our model fitted the data.

We observe that the confidence intervals of model IRRs overlap those computed from the data overlap in all instances. The empirical IRRs from the data also agree with previously noted patterns, e.g. for severe outcomes (hospitalisation or death) there is a very large increase in IRR for the oldest age group relative to the reference age group of 50–59-year-olds (Figure S1B). Similarly, we observe an increase in IRR as deprivation increases across all outcomes (Figure S1D). The results for ethnicity are mixed – non-White ethnicity categories had higher IRRs than the White category for the Pillar 2 positive test outcome, but lower IRRs than the White category for hospitalisation and death, which is because these IRRs are not adjusted for other covariates.

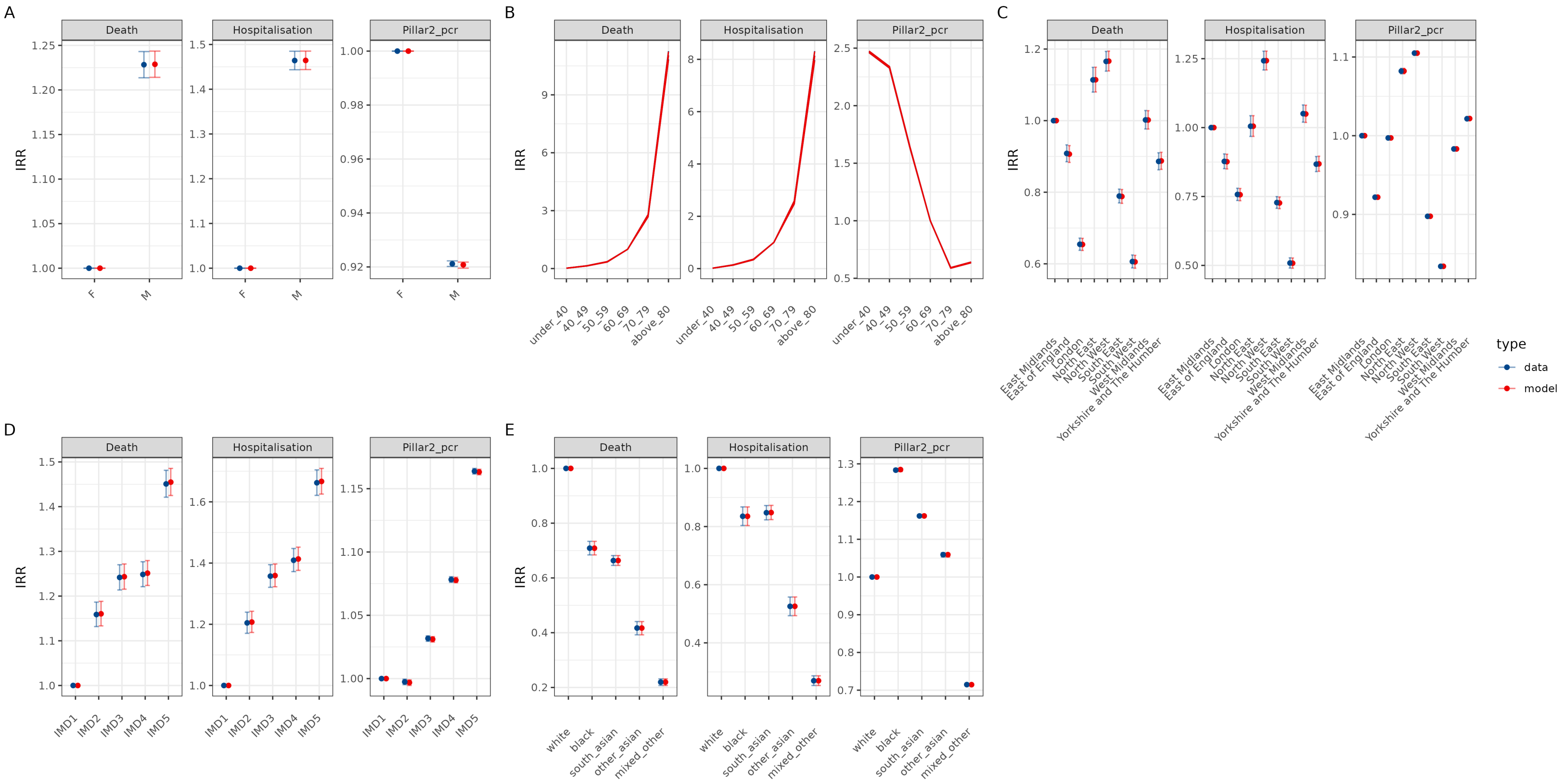

Figure S1: Marginal IRRs of data and fitted model for the full time period (May 2020-Feb 2021) for the preferred model). All reported IRRs are marginal and do not account for any other covariates.

#### A.2 Model selection

We parametrically captured variation by systematically examining evidence for interactions between covariates. We conducted model selection to identify the model which performed best across all three outcomes of interest (infection, hospitalisation, and deaths) by considering the average Akaike's information criterion (AIC) and coefficient of determination ($R^2$) across the three respective models and several different definitions for our covariates of interest.

We considered the following interactions:

- none
- Age and Region (A x R)
- Week and Age (W x A)
- Week and Region (W X R)
- Age and Region, Week and Age (A x R, W x A)
- Week and Region, Age and Region (W x R, A x R)
- Week and Region and Age (W x R x A)
- Week and Region, Week and Age (W x R, W x A)
- Age x Vaccination status (A x V)
- Week x Age, Age x Vaccination status (W x A, A x V)

For age we considered grouping Age by 10 year age bands and a simplified grouping which combined all ages below 40, 80 and above and used 10 year age bands in between. The definition of age is provided in section B.2

For ethnicity we considered

- 4 groups: White ethnicity, Black ethnicity, Asian ethnicity, Mixed or Other
- 6 groups: White ethnicity, Black ethnicity, South Asian ethnicity, Asian (other) ethnicity, Mixed, Other

For deprivation, we used the Index of Multiple Deprivation with either quintiles or deciles. Separately we provide our analysis for all sub-components of the IMD (with quintiles) in section E.

We run the model selection for the full time period and for each SARS-CoV-2 variant.

| Case definition | Time period | Age | Deprivation | Ethnicity | Interactions | RMSE | MAE | R2 | AIC |
|---|---|---|---|---|---|---|---|---|---|
| Death | Omicron | Age simplified | IMD quintiles | 5 groups | none | 0.28 | 0.05 | 0.55 | 38949.42 |
| Death | Wt | Age simplified | IMD quintiles | 5 groups | none | 1.53 | 0.30 | 0.79 | 52118.62 |
| Death | Alpha | Age simplified | IMD quintiles | 5 groups | none | 1.15 | 0.19 | 0.86 | 62811.70 |
| Death | Alpha | Age simplified | IMD quintiles | 5 groups | (AxR) | 1.70 | 0.22 | 0.69 | 66597.50 |
| Death | Alpha | Age simplified | IMD quintiles | 5 groups | (WxA) | 1.35 | 0.21 | 0.81 | 67421.76 |
| Death | Alpha | Age simplified | IMD quintiles | 5 groups | (AxR, WxA) | 1.67 | 0.22 | 0.70 | 71034.80 |
| Death | Delta | Age simplified | IMD quintiles | 5 groups | none | 0.20 | 0.03 | 0.33 | 85462.67 |
| Death | Delta | Age simplified | IMD quintiles | 5 groups | (AxR) | 0.21 | 0.04 | 0.28 | 86456.98 |
| Death | Full time period | Age simplified | IMD quintiles | 4 groups | none | 0.76 | 0.08 | 0.73 | 255633.34 |
| Death | Full time period | Age simplified | IMD quintiles | 5 groups | none | 0.72 | 0.07 | 0.74 | 256197.05 |
| Death | Full time period | Age simplified | IMD quintiles | 4 groups | (AxR) | 0.90 | 0.09 | 0.62 | 260966.16 |
| Death | Full time period | Age simplified | IMD quintiles | 5 groups | (AxR) | 0.85 | 0.08 | 0.64 | 261422.35 |
| Death | Full time period | Age (10yr bands) | IMD quintiles | 4 groups | none | 0.56 | 0.06 | 0.73 | 271085.90 |
| Death | Full time period | Age (10yr bands) | IMD quintiles | 5 groups | none | 0.54 | 0.05 | 0.73 | 271688.46 |
| Death | Full time period | Age (10yr bands) | IMD quintiles | 4 groups | (AxR) | 0.63 | 0.06 | 0.66 | 276167.02 |
| Death | Full time period | Age (10yr bands) | IMD quintiles | 5 groups | (AxR) | 0.61 | 0.06 | 0.65 | 277163.29 |
| Death | Full time period | Age simplified | IMD deciles | 4 groups | none | 0.52 | 0.06 | 0.69 | 303904.61 |
| Death | Full time period | Age simplified | IMD deciles | 5 groups | none | 0.50 | 0.05 | 0.69 | 304478.01 |
| Death | Full time period | Age simplified | IMD deciles | 4 groups | (AxR) | 0.59 | 0.06 | 0.60 | 309089.26 |
| Death | Full time period | Age simplified | IMD deciles | 5 groups | (AxR) | 0.57 | 0.06 | 0.59 | 309747.22 |
| Death | Full time period | Age (10yr bands) | IMD deciles | 4 groups | none | 0.39 | 0.04 | 0.68 | 321447.87 |
| Death | Full time period | Age (10yr bands) | IMD deciles | 5 groups | none | 0.37 | 0.04 | 0.68 | 321944.74 |
| Death | Full time period | Age (10yr bands) | IMD deciles | 4 groups | (AxR) | 0.43 | 0.04 | 0.61 | 326509.87 |
| Death | Full time period | Age (10yr bands) | IMD deciles | 5 groups | (AxR) | 0.41 | 0.04 | 0.61 | 326910.26 |
| Death | Full time period | Age simplified | IMD deciles | 4 groups | (WxR) | 0.78 | 0.06 | 0.30 | 327965.22 |
| Death | Full time period | Age simplified | IMD deciles | 4 groups | (WxRxA) | 0.78 | 0.06 | 0.30 | 327965.22 |
| Death | Full time period | Age (10yr bands) | IMD deciles | 5 groups | (WxR, AxR) | 0.63 | 0.04 | 0.10 | 354884.73 |

Figure S2: Top models for death, ranked by AIC.

| Case definition | Time period | Age | Deprivation | Ethnicity | Interactions | RMSE | MAE | R2 | AIC |
|---|---|---|---|---|---|---|---|---|---|
| Death | Omicron | Age simplified | IMD quintiles | 5 groups | none | 0.28 | 0.05 | 0.55 | 38949.42 |
| Death | Wt | Age simplified | IMD quintiles | 5 groups | none | 1.53 | 0.30 | 0.79 | 52118.62 |
| Death | Alpha | Age simplified | IMD quintiles | 5 groups | none | 1.15 | 0.19 | 0.86 | 62811.70 |
| Death | Alpha | Age simplified | IMD quintiles | 5 groups | (AxR) | 1.70 | 0.22 | 0.69 | 66597.50 |
| Death | Alpha | Age simplified | IMD quintiles | 5 groups | (WxA) | 1.35 | 0.21 | 0.81 | 67421.76 |
| Death | Alpha | Age simplified | IMD quintiles | 5 groups | (AxR, WxA) | 1.67 | 0.22 | 0.70 | 71034.80 |
| Death | Delta | Age simplified | IMD quintiles | 5 groups | none | 0.20 | 0.03 | 0.33 | 85462.67 |
| Death | Delta | Age simplified | IMD quintiles | 5 groups | (AxR) | 0.21 | 0.04 | 0.28 | 86456.98 |
| Death | Full time period | Age simplified | IMD quintiles | 4 groups | none | 0.76 | 0.08 | 0.73 | 255633.34 |
| Death | Full time period | Age simplified | IMD quintiles | 5 groups | none | 0.72 | 0.07 | 0.74 | 256197.05 |
| Death | Full time period | Age simplified | IMD quintiles | 4 groups | (AxR) | 0.90 | 0.09 | 0.62 | 260966.16 |
| Death | Full time period | Age simplified | IMD quintiles | 5 groups | (AxR) | 0.85 | 0.08 | 0.64 | 261422.35 |
| Death | Full time period | Age (10yr bands) | IMD quintiles | 4 groups | none | 0.56 | 0.06 | 0.73 | 271085.90 |
| Death | Full time period | Age (10yr bands) | IMD quintiles | 5 groups | none | 0.54 | 0.05 | 0.73 | 271688.46 |
| Death | Full time period | Age (10yr bands) | IMD quintiles | 4 groups | (AxR) | 0.63 | 0.06 | 0.66 | 276167.02 |
| Death | Full time period | Age (10yr bands) | IMD quintiles | 5 groups | (AxR) | 0.61 | 0.06 | 0.65 | 277163.29 |
| Death | Full time period | Age simplified | IMD deciles | 4 groups | none | 0.52 | 0.06 | 0.69 | 303904.61 |
| Death | Full time period | Age simplified | IMD deciles | 5 groups | none | 0.50 | 0.05 | 0.69 | 304478.01 |
| Death | Full time period | Age simplified | IMD deciles | 4 groups | (AxR) | 0.59 | 0.06 | 0.60 | 309089.26 |
| Death | Full time period | Age simplified | IMD deciles | 5 groups | (AxR) | 0.57 | 0.06 | 0.59 | 309747.22 |
| Death | Full time period | Age (10yr bands) | IMD deciles | 4 groups | none | 0.39 | 0.04 | 0.68 | 321447.87 |
| Death | Full time period | Age (10yr bands) | IMD deciles | 5 groups | none | 0.37 | 0.04 | 0.68 | 321944.74 |
| Death | Full time period | Age (10yr bands) | IMD deciles | 4 groups | (AxR) | 0.43 | 0.04 | 0.61 | 326509.87 |
| Death | Full time period | Age (10yr bands) | IMD deciles | 5 groups | (AxR) | 0.41 | 0.04 | 0.61 | 326910.26 |
| Death | Full time period | Age simplified | IMD deciles | 4 groups | (WxR) | 0.78 | 0.06 | 0.30 | 327965.22 |
| Death | Full time period | Age simplified | IMD deciles | 4 groups | (WxRxA) | 0.78 | 0.06 | 0.30 | 327965.22 |
| Death | Full time period | Age (10yr bands) | IMD deciles | 5 groups | (WxR, AxR) | 0.63 | 0.04 | 0.10 | 354884.73 |

Figure S3: Top models for hospitalisation, ranked by AIC.

| Case definition | Time period | Age | Deprivation | Ethnicity | Interactions | RMSE | MAE | R2 | AIC |
|---|---|---|---|---|---|---|---|---|---|
| Death | Omicron | Age simplified | IMD quintiles | 5 groups | none | 0.28 | 0.05 | 0.55 | 38949.42 |
| Death | Wt | Age simplified | IMD quintiles | 5 groups | none | 1.53 | 0.30 | 0.79 | 52118.62 |
| Death | Alpha | Age simplified | IMD quintiles | 5 groups | none | 1.15 | 0.19 | 0.86 | 62811.70 |
| Death | Alpha | Age simplified | IMD quintiles | 5 groups | (AxR) | 1.70 | 0.22 | 0.69 | 66597.50 |
| Death | Alpha | Age simplified | IMD quintiles | 5 groups | (WxA) | 1.35 | 0.21 | 0.81 | 67421.76 |
| Death | Alpha | Age simplified | IMD quintiles | 5 groups | (AxR, WxA) | 1.67 | 0.22 | 0.70 | 71034.80 |
| Death | Delta | Age simplified | IMD quintiles | 5 groups | none | 0.20 | 0.03 | 0.33 | 85462.67 |
| Death | Delta | Age simplified | IMD quintiles | 5 groups | (AxR) | 0.21 | 0.04 | 0.28 | 86456.98 |
| Death | Full time period | Age simplified | IMD quintiles | 4 groups | none | 0.76 | 0.08 | 0.73 | 255633.34 |
| Death | Full time period | Age simplified | IMD quintiles | 5 groups | none | 0.72 | 0.07 | 0.74 | 256197.05 |
| Death | Full time period | Age simplified | IMD quintiles | 4 groups | (AxR) | 0.90 | 0.09 | 0.62 | 260966.16 |
| Death | Full time period | Age simplified | IMD quintiles | 5 groups | (AxR) | 0.85 | 0.08 | 0.64 | 261422.35 |
| Death | Full time period | Age (10yr bands) | IMD quintiles | 4 groups | none | 0.56 | 0.06 | 0.73 | 271085.90 |
| Death | Full time period | Age (10yr bands) | IMD quintiles | 5 groups | none | 0.54 | 0.05 | 0.73 | 271688.46 |
| Death | Full time period | Age (10yr bands) | IMD quintiles | 4 groups | (AxR) | 0.63 | 0.06 | 0.66 | 276167.02 |
| Death | Full time period | Age (10yr bands) | IMD quintiles | 5 groups | (AxR) | 0.61 | 0.06 | 0.65 | 277163.29 |
| Death | Full time period | Age simplified | IMD deciles | 4 groups | none | 0.52 | 0.06 | 0.69 | 303904.61 |
| Death | Full time period | Age simplified | IMD deciles | 5 groups | none | 0.50 | 0.05 | 0.69 | 304478.01 |
| Death | Full time period | Age simplified | IMD deciles | 4 groups | (AxR) | 0.59 | 0.06 | 0.60 | 309089.26 |
| Death | Full time period | Age simplified | IMD deciles | 5 groups | (AxR) | 0.57 | 0.06 | 0.59 | 309747.22 |
| Death | Full time period | Age (10yr bands) | IMD deciles | 4 groups | none | 0.39 | 0.04 | 0.68 | 321447.87 |
| Death | Full time period | Age (10yr bands) | IMD deciles | 5 groups | none | 0.37 | 0.04 | 0.68 | 321944.74 |
| Death | Full time period | Age (10yr bands) | IMD deciles | 4 groups | (AxR) | 0.43 | 0.04 | 0.61 | 326509.87 |
| Death | Full time period | Age (10yr bands) | IMD deciles | 5 groups | (AxR) | 0.41 | 0.04 | 0.61 | 326910.26 |
| Death | Full time period | Age simplified | IMD deciles | 4 groups | (WxR) | 0.78 | 0.06 | 0.30 | 327965.22 |
| Death | Full time period | Age simplified | IMD deciles | 4 groups | (WxRxA) | 0.78 | 0.06 | 0.30 | 327965.22 |
| Death | Full time period | Age (10yr bands) | IMD deciles | 5 groups | (WxR, AxR) | 0.63 | 0.04 | 0.10 | 354884.73 |

Figure S4: Top models for Pillar 2 PCR positive cases, ranked by AIC.

### B Data description

#### B.1 Ethnicity

Ethnicity data is obtained from three sources: the case database, NIMS database, and the ONS Census 2021 data [3]. The challenge is that there is no consistent definition for ethnicity.

We created the following mapping for reported fields (in case database and NIMS database) to a simplified list of ethnicities:

- **Unknown ethnicity**: 'Prefer not to say', 'c19ds HES - unable to match'
- **White ethnicity**: 'British, Mixed British', 'British (White)', 'Irish', 'Irish (White)', 'Any other White background'
- **Mixed ethnicity**: 'White and Asian', 'White and Black African'
- **Other ethnicity**: 'Any other mixed background', 'Any other ethnic group'
- **Indian ethnicity**: 'Indian or British Indian', 'Indian (Asian or Asian British)'
- **Pakistani or Bangladeshi ethnicity**: 'Bangladeshi or British Bangladeshi', 'Bangladeshi (Asian or Asian British)'
- **Other Asian ethnicity**: 'Any other Asian background', 'Chinese', 'Chinese (other ethnic group)'
- **Black ethnicity**: 'African', 'Any other Black background', 'Caribbean', 'African (Black or Black British)', 'White and Black African (Mixed)', 'Caribbean (Black or Black British)'

The mapping was constructed so that the resultant groupings approximately matched the Census 2021 data for the proportion of individuals in the case and NIMS database. For example 'British, Mixed British' is a large group bigger than the total number of people classified as Mixed ethnicity for England combined. Similarly we chose to include 'White and Black African (Mixed)' with Black ethnicity rather than Mixed ethnicity.

Using this mapping we could create to sets of ethnicity categories:

1. 4 ethnicities: White ethnicity, Black ethnicity, Asian ethnicity, and Mixed/Other
2. 5 ethnicities: White ethnicity, Black ethnicity, South Asian ethnicity, Asian (other) ethnicity, and Mixed/ Other

The Census 2021 database allowed us to create datasets with the number of individuals for each LTLA by sex (male/female), age (1-100 in 1 year increments) and ethnicity. For 292 out of 306 English LTLAs we can obtain this breakdown with an ethnicity granularity of 20 ethnicities. For the remaining 14 LTLAs we can only obtain the data with an ethnicity granularity of 6 ethnicities (due to the fact that some of the groups have too few individuals in).

We remapped the ethnicities reported in the ONS Census data to our 6 ethnicities grouping (and separately to the 4 ethnicities grouping). For the 14 LTLAs for which we only had 6 ethnicity granularity we split Asian ethnicity into South Asian and Asian (other) according to the national proportion of the two (74.51% of the Asian population of England are South Asian).

This data was used to assign the 'unknown ethnicity' category by using randomly generated uniform variables between zero and one. For each individual with 'unknown ethnicity' we compared the random number vs the cumulative ethnicity distribution for each (LTLA, sex, age, ethnicity) combination. That is if 70% of individuals were of White ethnicity, 10% of Black ethnicity, 10% of South Asian Ethnicity, 5% of Asian (other) ethnicity, 5% Mixed/Other ethnicity, the cumulative distribution would be 70%, 80%, 90%, 95%, 100% and a uniform random number between 0 and less than 0.7 would be assigned to White ethnicity, between 0.7 and less than 0.8 to Black ethnicity etc.

The case and NIMS database only cover individuals which had an infection or received at least one vaccination. This did not cover all of the population of England as non vaccinated individuals who had no infection are missing. The largest group for this are the under 18s and in particular the under 12s

| **Ethnicity** | **n** | **Percentage** |
|---|---:|---:|
| Black | 2,381,137 | 4.2% |
| Mixed/Other | 2,896,718 | 5.2% |
| Asian (other) | 1,379,484 | 2.4% |
| South Asian | 4,044,784 | 7.2% |
| White | 45,725,274 | 81.0% |
| *Total English population* | *56,427,397* | |

Table S2: Number and percentage of English population by ethnicity (ONS Census 2021)

(as vaccination in that group was less common). We create entries in our dataset for all the missing individuals using the numbers of people in each LTLA, sex, age, ethnicity group from the ONS Census 2021 data.

For the overall English population we provide a breakdown by ethnicity in Table S2.

#### B.2 Computation of age

Integer age in years at the time of an event (e.g. positive test, vaccination, or death) was available from both the case database and NIMS database. To obtain a single consistent age for each individual, we imputed age for every individual in the population on 1st January 2020 using the mid-point of the precise age from within the range consistent with the event-specific ages available for each individual.

$$\begin{aligned} min\_birthday &= \max_{i \in events} date_i - (age_i + 1) \\ max\_birthday &= \min_{i \in events} date_i - age_i \\ birthday &= \frac{min\_birthday + max\_birthday}{2} \end{aligned}$$

Using this estimated birthday we calculated the age of the individual on 1 January 2020.

#### B.3 Vaccination

Vaccination data was available from the NIMS database. From this data we computed for each week how many people were in one of the following categories:

- not vaccinated
- First dose $<$ 21 days since vaccination
- First dose $>$ 21 days since vaccination
- Second dose $<$ 14 days since vaccination
- Second dose $>$ 14 days since vaccination
- Waning of vaccination: 10-18 weeks since vaccination
- Waning of vaccination: 18+ weeks since vaccination

We also know what vaccine each individual received and we grouped these into the following:

- mRNA: Pfizer and Moderna
- Adenovirus: AZ and Janssen
- Mixed dose (if first and second vaccine were different)
- Novavax
- Unknown vaccine product

#### B.4 Index of Multiple Deprivation

Deprivation has been measured by the Local Communities Department of the UK Government since the 1970s at spatially fine scale. The data is typically published every 4 years and the current version is the English Indices of Deprivation 2019 (IoD2019, see below schematic).

The official measure of relative deprivation in England is the Index of Multiple Deprivation (IMD), which is part of the IoD2019. A broad range of indicators and data is used to compute the 7 domains of the IMD (see list below) and the overall IMD is a weighted average of those domains (methodology and further details see [4]).

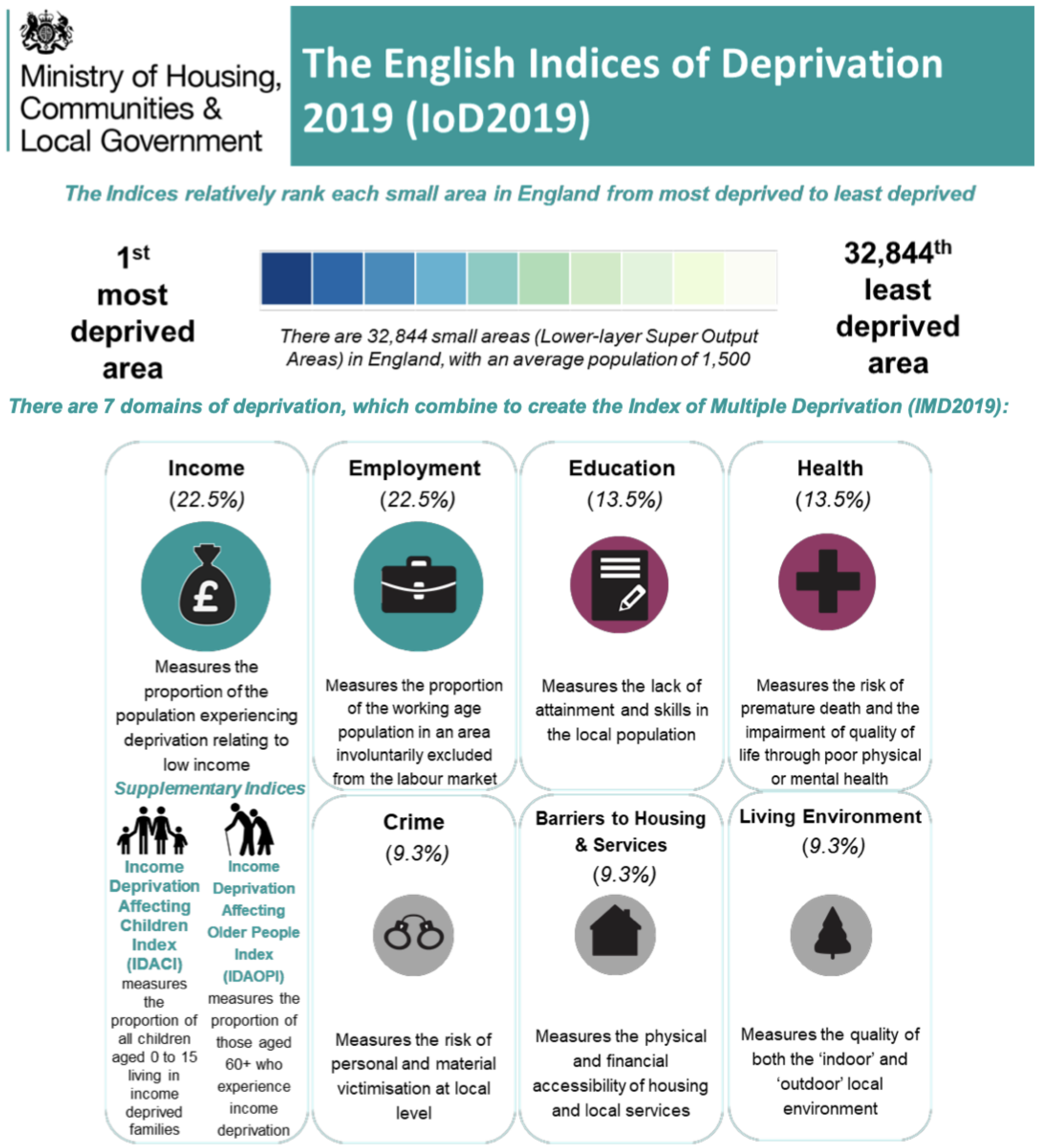

Components of the Index of Multiple Deprivation (IMD) published by the UK Government [4]

There are 39 indicators, which are combined into seven domains. The overall IMD is the weighted average of the seven domains and is reported at the Lower-layer Super Output Area (LSOA) level (these correspond to neighbourhoods). The score is ranked across all LSOAs, with high ranks corresponding to the most deprived areas. The IMD measure is strictly a rank.

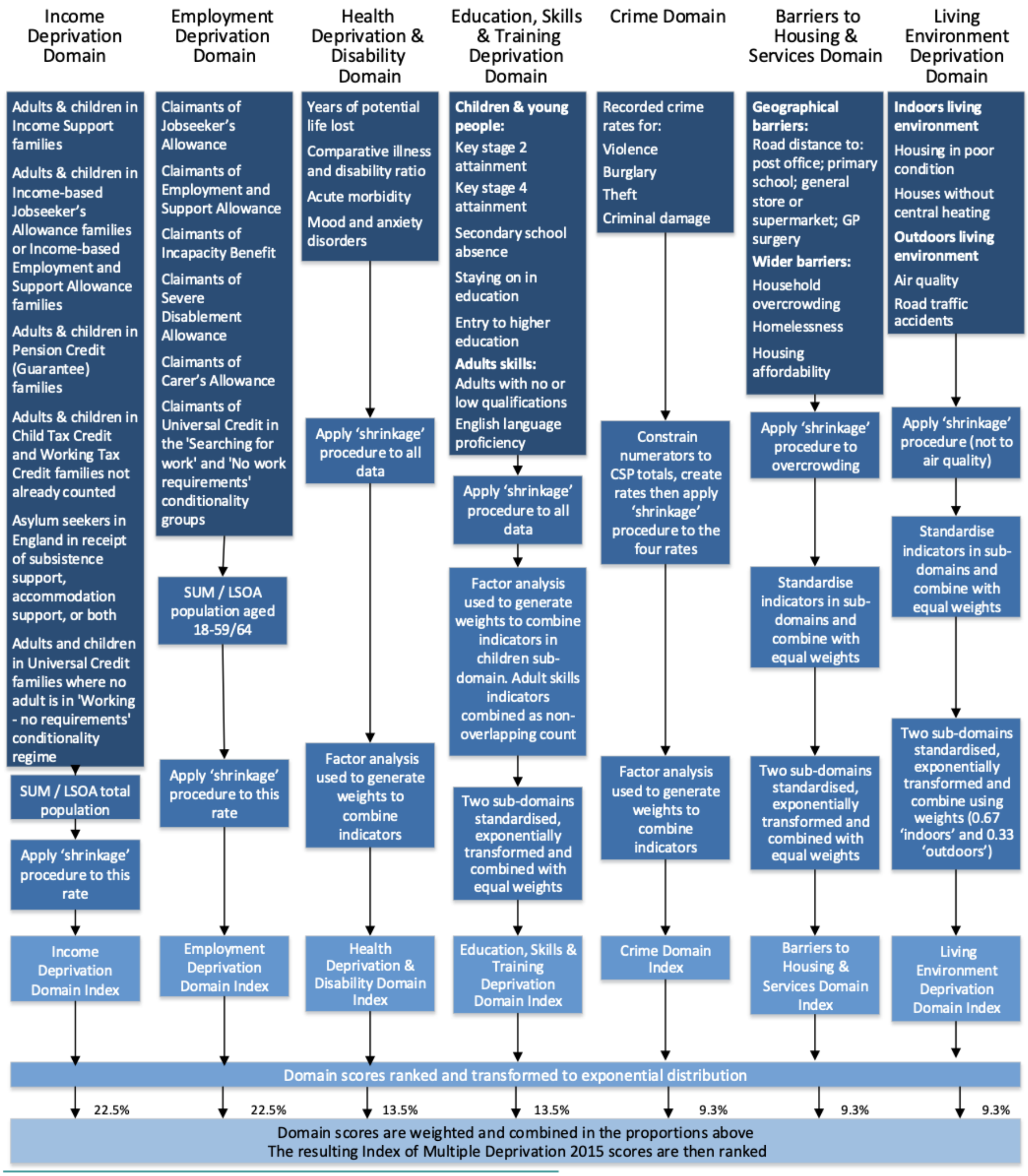

Summary of the domains, indicators and data used to create the Indices of Deprivation 2019 [4]

The IMD ranks are available at the LSOA level, however they can be aggregated into larger area units. In our work we use the aggregated data to LTLA level.

### C Information on time periods

#### C.1 Dominant Variant by region

We determined the dominant variant using S-target positive and negative data from the case database and consider the average by week and region. Figure S5 shows plots the dominant variant by week and region.

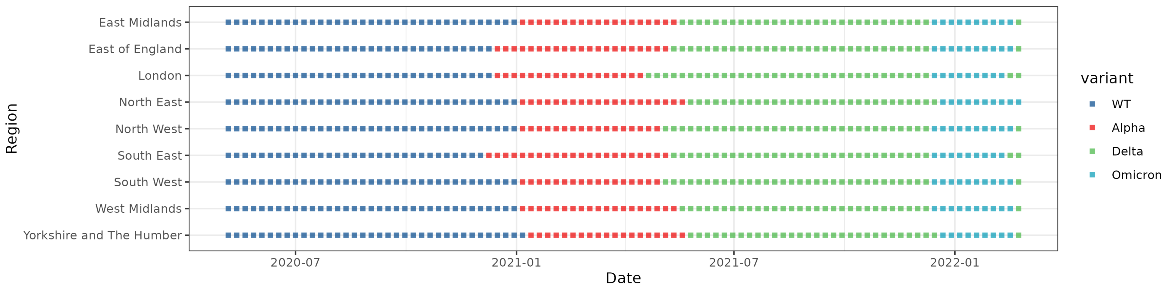

Figure S5: Dominant SARS-CoV-2 variant by region and week

#### C.2 Average contact rates

We use the Imperial College YouGov Covid Behavioural tracker study to [5] to estimate the average number of contacts. This is based on repeated cross-sectional surveys which we re-aggregated into our age grouping.

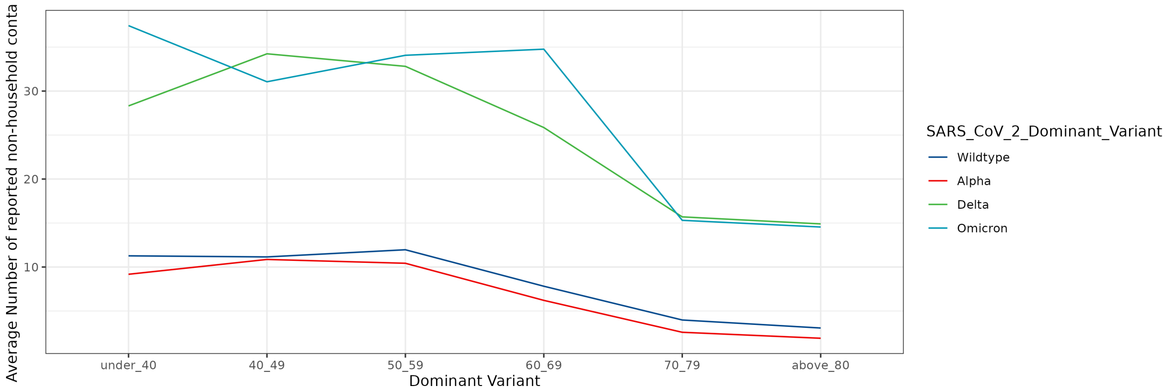

Figure S6: Average contact rate by age group and dominant SARS-CoV-2 variant

#### C.3 Definition of restriction periods & NPIs for England

We used ONS data (up to December 2020) which contained LTLA-level restrictions according to Tiers which were in use in the second half of 2020. We also have information on non-pharmaceutical interventions reported in the Oxford Covid Government Response Tracker [6, 7] (see Figure S8 which displays the combined stringency index as well as Containment and closure policies, Health Systems policies, and Economic policies).

We used the Containment and closure policy measures to construct the restriction levels in Figure S7 as follows:

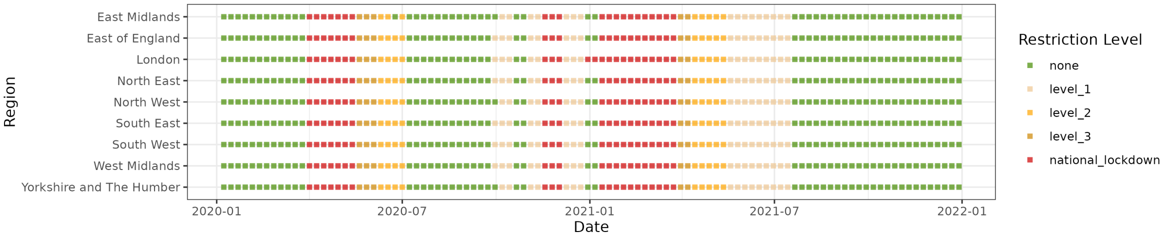

Figure S7: Restriction level by region and week.

- C6=2 (maximum score for stay-at-home requirement) implies a **national lockdown**
- If C6<2 and we have an ONS tier restriction we use the level of the tier restriction
- We use **national lockdown** for isoweek 52 and 53 (2020) the London region (Christmas 2020 lockdown as London was in tier 4)
- **Level 3** for isoweek 12 and 13 (2021) [Roadmap phase 1]
- **Level 2** for isoweek 14 to 18 (2021) [Roadmap phase 2]
- If C6<2, no ONS restriction is in place, C4=4 (gatherings restricted to 10 people or less) and C1<2 (some school restrictions recommended but no closing) implies **Level 1**
- If C6<2, no ONS restriction is in place, C4=4 (gatherings restricted to 10 people or less) and C1=2 (some school closing recommended) implies **Level 2**
- otherwise the restriction level is set to **none**.

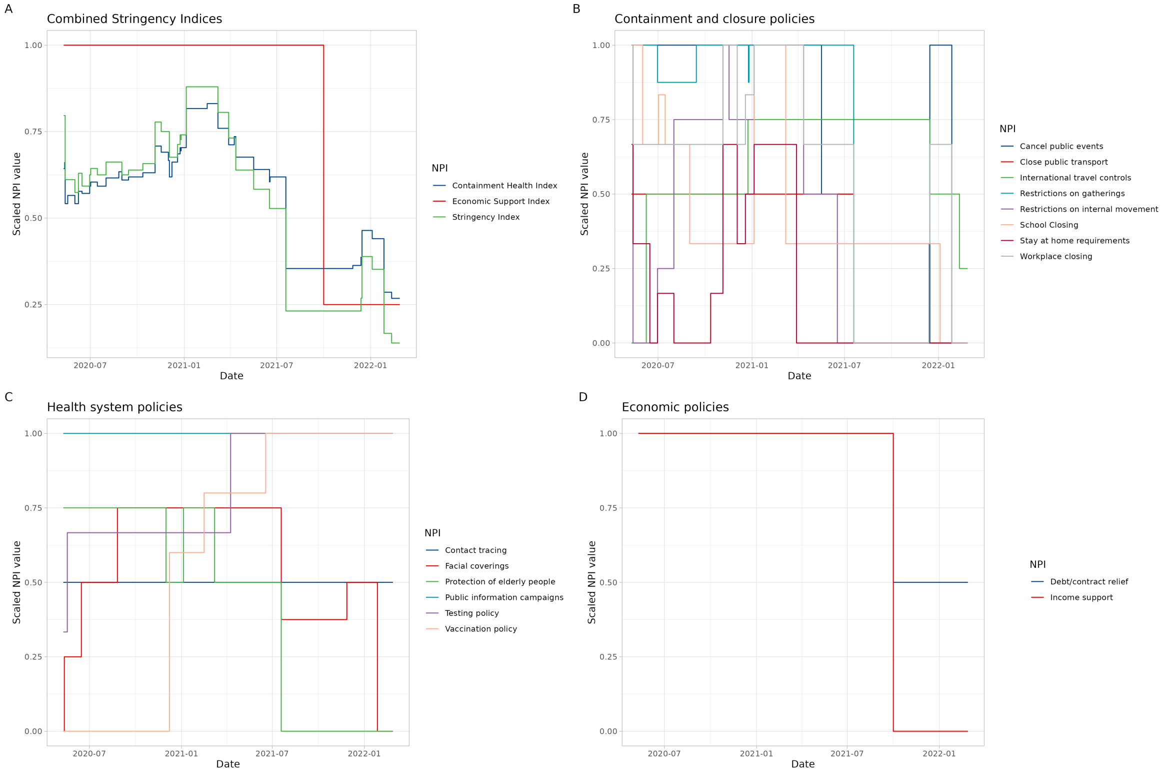

Figure S8: Non-pharmaceutical interventions in England for study period (Oxford Covid Government Response Tracker [7, 6])

We assign the following (linearly increasing) numeric values to each restriction level:

- None = 1
- Level 1 = 2
- Level 2 = 3
- Level 3 = 4
- National lockdown = 5

#### C.4 Self-isolation rules

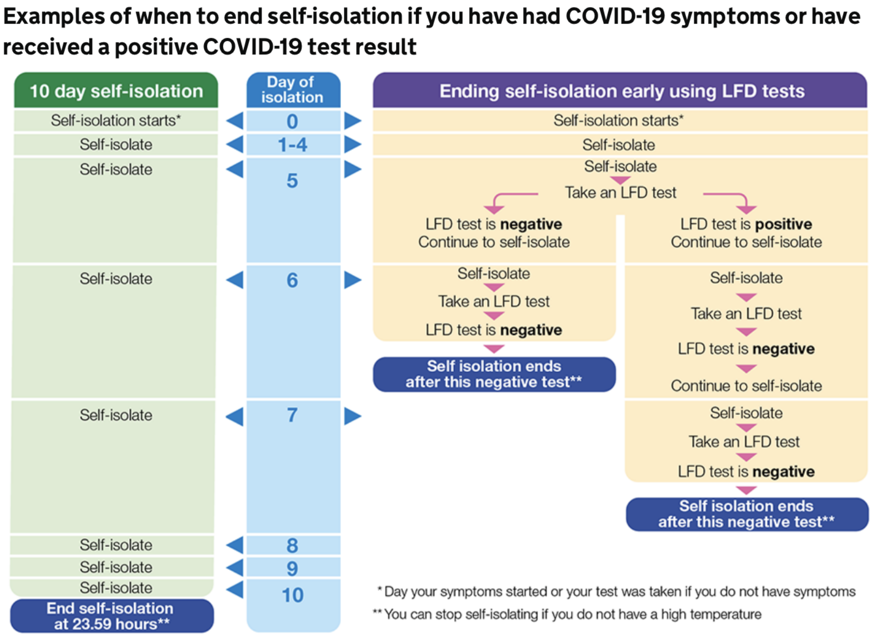

Figure S9: Stay at home: guidance for households with possible or confirmed COVID-19 infection

### D Additional results figures

| Variable | Category | n | Person days at risk | Cases (Pillar 2 PCR) | Hospitalisations | Deaths |
|---|---|---|---|---|---|---|
| Vaccination status | Not Vaccinated | 12,524,706 | 21,652,863,421 | 7,030,900 | 289,451 | 80,601 |
| | First Dose <21d Adenovirus | 454 | 422,148,818 | 31,282 | 6,179 | 2,565 |
| | First Dose <21d mRNA | 253,760 | 438,532,528 | 210,475 | 7,628 | 2,364 |
| | First Dose <21d Novavax | 13 | 861 | 4 | - | - |
| | First Dose <21d Unknown | 483 | 630,189 | 1,024 | 47 | 6 |
| | First Dose >3w Adenovirus | 343,341 | 1,168,792,975 | 97,771 | 6,782 | 2,419 |
| | First Dose >3w mRNA | 2,462,228 | 1,187,931,423 | 542,990 | 8,482 | 2,388 |
| | First Dose >3w Novavax | 16 | 3,360 | 20 | - | - |
| | First Dose >3w Unknown | 4,636 | 1,912,680 | 2,068 | 50 | 4 |
| | Second Dose <2w Adenovirus | 1,149 | 197,762,544 | 25,825 | 292 | 12 |
| | Second Dose <2w Mixed Dose | 6,699 | 1,618,484 | 1,855 | 38 | 2 |
| | Second Dose <2w mRNA | 331,407 | 234,175,466 | 47,748 | 627 | 66 |
| | Second Dose <2w Novavax | 1 | 2,233 | 13 | - | - |
| | Second Dose <2w Unknown | 192 | 153,762 | 173 | 4 | - |
| | Second Dose >2w Adenovirus | 19,049 | 842,430,442 | 172,385 | 2,224 | 160 |
| | Second Dose >2w Mixed Dose | 83,543 | 6,804,343 | 4,089 | 135 | 16 |
| | Second Dose >2w mRNA | 1,206,396 | 829,053,400 | 113,626 | 1,686 | 132 |
| | Second Dose >2w Novavax | 11 | 42,385 | 79 | 2 | - |
| | Second Dose >2w Unknown | 1,421 | 991,018 | 762 | 4 | - |
| | Second Dose 10-18w Adenovirus | 42,560 | 1,063,198,297 | 362,165 | 8,399 | 1,008 |
| | Second Dose 10-18w Mixed Dose | 58,654 | 3,697,260 | 2,450 | 143 | 32 |
| | Second Dose 10-18w mRNA | 967,000 | 919,526,650 | 373,924 | 4,419 | 398 |
| | Second Dose 10-18w Novavax | - | 40,124 | 68 | 1 | - |
| | Second Dose 10-18w Unknown | 1,388 | 1,561,420 | 1,878 | 30 | - |
| | Second Dose over 18w Adenovirus | 2,781,388 | 1,561,139,076 | 1,068,220 | 33,061 | 5,879 |
| | Second Dose over 18w Mixed Dose | 24,280 | 4,203,626 | 3,511 | 205 | 63 |
| | Second Dose over 18w mRNA | 4,066,262 | 1,129,558,164 | 751,049 | 18,615 | 3,765 |
| | Second Dose over 18w Novavax | 527 | 82,565 | 264 | 1 | - |
| | Second Dose over 18w Unknown | 9,527 | 2,761,353 | 5,248 | 166 | 3 |
| | Booster 10-18w Adenovirus | 170,587 | 225,869,399 | 194,668 | 2,385 | 188 |
| | Booster 10-18w mRNA | 301,068 | 178,424,050 | 168,635 | 1,738 | 151 |
| | Booster 10-18w Novavax | 151 | 3,906 | 37 | - | - |
| | Booster 10-18w Unknown | 452 | 281,008 | 774 | 8 | - |
| | Booster Dose <2w Adenovirus | 7,434,225 | 772,914,618 | 428,088 | 10,506 | 966 |
| | Booster Dose <2w mRNA | 6,054,894 | 586,137,083 | 294,073 | 7,356 | 770 |
| | Booster Dose <2w Novavax | 208 | 10,528 | 37 | - | - |
| | Booster Dose <2w Unknown | 7,776 | 1,040,018 | 2,320 | 54 | - |
| | Booster Dose >2w Adenovirus | 9,188,109 | 332,906,007 | 137,555 | 11,487 | 1,522 |
| | Booster Dose >2w mRNA | 5,512,367 | 326,242,322 | 207,494 | 14,032 | 1,809 |
| | Booster Dose >2w Novavax | 128 | 5,992 | 36 | - | - |
| | Booster Dose >2w Unknown | 16,146 | 803,922 | 2,482 | 118 | 1 |
| | Booster over 18w Adenovirus | 508,392 | 12,907,790 | 5,238 | 661 | 148 |
| | Booster over 18w Mixed Dose | 7 | 133 | 4 | - | - |
| | Booster over 18w mRNA | 1,954,193 | 43,244,152 | 16,974 | 2,059 | 385 |
| | Booster over 18w Novavax | 146 | 4,984 | 28 | - | - |
| | Booster over 18w Unknown | 4,470 | 102,228 | 176 | 8 | - |

Figure S10: Breakdown of synthetic population by vaccination status (n is the count of individuals in this category at the end of the study period, person days at risk and counts of Pillar 2 PCR positive cases, hospitalisations and deaths are over full study period - as a result the number of cases may be higher than the final number of individuals in that category).

Figure S11 provides the IRRs for covariates which are not plotted in Figure 3 of the main text.

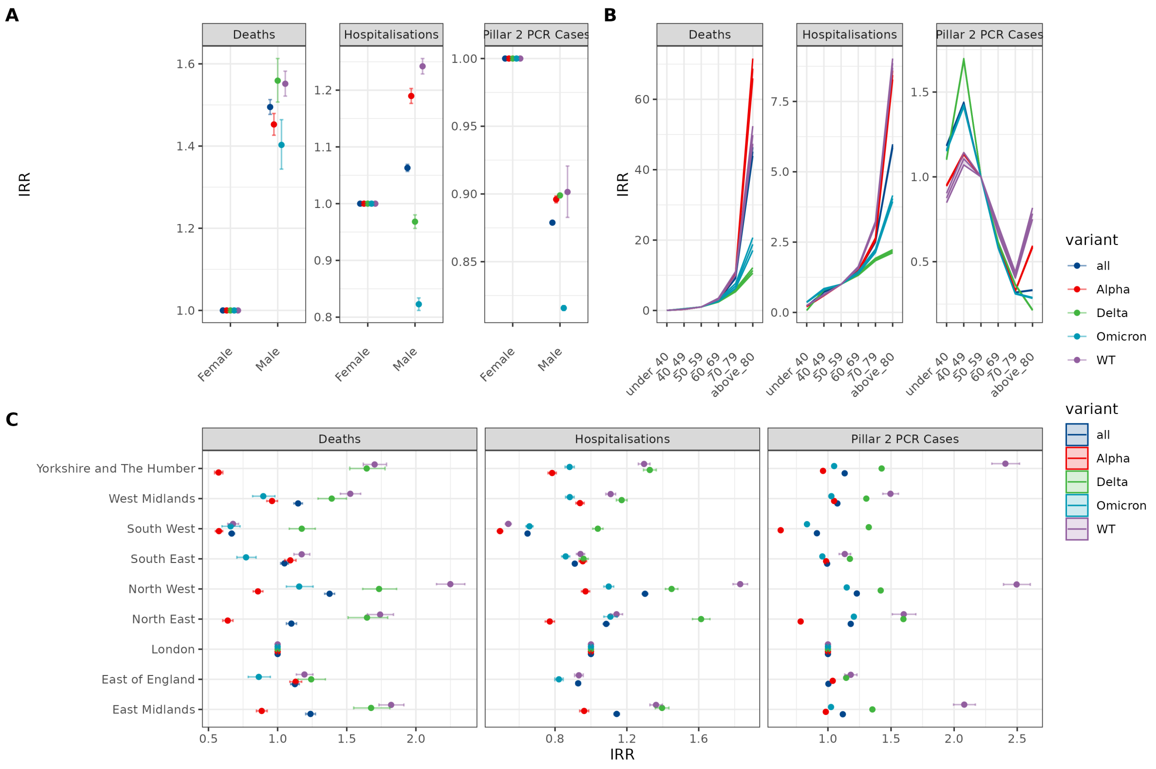

Figure S11: Estimated IRRs for (A) sex, (B) age, and (c) region covariates from preferred models. Age was categorised by under 40, 10 year age band up to 79 and over 80s, with 50-59 being the reference group (IRR=1). Results are shown for the whole pandemic ('all' - test dates between May 2020 and February 2022) and for the time intervals within that period where specific viral variants dominated (see SI for details). WT=Wild-Type (pre-December 2020).

Table S3 provides the IRRs for vaccine status and the associated VE estimates, including 95% confidence intervals.

| Vaccine | IRR (Deaths) | IRR (Hospitalisations) | IRR (Pillar 2 PCR Cases) | Vaccine effectiveness (Deaths) | Vaccine effectiveness (Hospitalisations) | Vaccine effectiveness (Pillar 2 PCR Cases) |
|---|---|---|---|---|---|---|
| **Full time period** | | | | | | |
| Not Vaccinated | 1 (1-1) | 1 (1-1) | 1 (1-1) | 0 (0-0) | 0 (0-0) | 0 (0-0) |
| Less than 3w after first dose Adenovirus | 0.297 (0.284-0.309) | 0.701 (0.682-0.72) | 0.924 (0.913-0.934) | 0.703 (0.691-0.716) | 0.299 (0.28-0.318) | 0.076 (0.066-0.087) |
| Less than 3w after first dose mRNA | 0.161 (0.155-0.168) | 0.444 (0.433-0.454) | 0.869 (0.865-0.872) | 0.839 (0.832-0.845) | 0.556 (0.546-0.567) | 0.131 (0.128-0.135) |
| Over 3w after first dose Adenovirus | 0.276 (0.263-0.289) | 0.274 (0.267-0.281) | 0.407 (0.404-0.41) | 0.724 (0.711-0.737) | 0.726 (0.719-0.733) | 0.593 (0.59-0.596) |
| Over 3w after first dose mRNA | 0.152 (0.145-0.159) | 0.24 (0.235-0.246) | 0.514 (0.513-0.516) | 0.848 (0.841-0.855) | 0.76 (0.754-0.765) | 0.486 (0.484-0.487) |
| Less than 2w after second dose Adenovirus | 0.053 (0.03-0.095) | 0.098 (0.087-0.11) | 0.359 (0.355-0.364) | 0.947 (0.905-0.97) | 0.902 (0.89-0.913) | 0.641 (0.636-0.645) |
| Less than 2w after second dose mRNA | 0.038 (0.03-0.049) | 0.121 (0.112-0.131) | 0.244 (0.242-0.246) | 0.962 (0.951-0.97) | 0.879 (0.869-0.888) | 0.756 (0.754-0.758) |
| Less than 2w after second dose Mixed Dose | .. | .. | 0.379 (0.362-0.397) | .. | .. | 0.621 (0.603-0.638) |
| 2-10w after second dose Adenovirus | 0.087 (0.074-0.103) | 0.075 (0.071-0.078) | 0.315 (0.314-0.317) | 0.913 (0.897-0.926) | 0.925 (0.922-0.929) | 0.685 (0.683-0.686) |
| 2-10w after second dose Mixed Dose | 0.144 (0.088-0.236) | 0.109 (0.092-0.129) | 0.273 (0.265-0.281) | 0.856 (0.764-0.912) | 0.891 (0.871-0.908) | 0.727 (0.719-0.735) |
| 2-10w after second dose mRNA | 0.033 (0.028-0.039) | 0.089 (0.085-0.094) | 0.155 (0.154-0.156) | 0.967 (0.961-0.972) | 0.911 (0.906-0.915) | 0.845 (0.844-0.846) |
| 10-18w after second dose Adenovirus | 0.129 (0.12-0.139) | 0.116 (0.113-0.119) | 0.42 (0.418-0.421) | 0.871 (0.861-0.88) | 0.884 (0.881-0.887) | 0.58 (0.579-0.582) |
| 10-18w after second dose Mixed Dose | 0.309 (0.218-0.438) | 0.229 (0.194-0.27) | 0.466 (0.448-0.484) | 0.691 (0.562-0.782) | 0.771 (0.73-0.806) | 0.534 (0.516-0.552) |
| 10-18w after second dose mRNA | 0.062 (0.056-0.069) | 0.086 (0.083-0.088) | 0.331 (0.33-0.333) | 0.938 (0.931-0.944) | 0.914 (0.912-0.917) | 0.669 (0.667-0.67) |
| Over 18w after second dose Adenovirus | 0.228 (0.22-0.238) | 0.215 (0.212-0.218) | 0.514 (0.513-0.516) | 0.772 (0.762-0.78) | 0.785 (0.782-0.788) | 0.486 (0.484-0.487) |
| Over 18w after second dose Mixed Dose | 0.265 (0.207-0.34) | 0.241 (0.21-0.277) | 0.53 (0.513-0.548) | 0.735 (0.66-0.793) | 0.759 (0.723-0.79) | 0.47 (0.452-0.487) |
| Over 18w after second dose mRNA | 0.132 (0.127-0.138) | 0.152 (0.15-0.155) | 0.487 (0.486-0.489) | 0.868 (0.862-0.873) | 0.848 (0.845-0.85) | 0.513 (0.511-0.514) |
| Less than 2w after booster dose Adenovirus | 0.041 (0.035-0.047) | 0.074 (0.071-0.077) | 0.332 (0.331-0.334) | 0.959 (0.953-0.965) | 0.926 (0.923-0.929) | 0.668 (0.666-0.669) |
| Less than 2w after booster dose mRNA | 0.029 (0.024-0.034) | 0.08 (0.076-0.084) | 0.382 (0.38-0.384) | 0.971 (0.966-0.976) | 0.92 (0.916-0.924) | 0.618 (0.616-0.62) |
| 2-10w after booster dose Adenovirus | 0.045 (0.042-0.048) | 0.059 (0.057-0.06) | 0.249 (0.249-0.25) | 0.955 (0.952-0.958) | 0.941 (0.94-0.943) | 0.751 (0.75-0.751) |
| 2-10w after booster dose mRNA | 0.03 (0.028-0.032) | 0.067 (0.065-0.069) | 0.284 (0.283-0.285) | 0.97 (0.968-0.972) | 0.933 (0.931-0.935) | 0.716 (0.715-0.717) |
| 10-18w after booster dose Adenovirus | 0.072 (0.068-0.077) | 0.095 (0.093-0.097) | 0.392 (0.39-0.395) | 0.928 (0.923-0.932) | 0.905 (0.903-0.907) | 0.608 (0.605-0.61) |
| 10-18w after booster dose mRNA | 0.046 (0.043-0.049) | 0.083 (0.081-0.085) | 0.437 (0.435-0.439) | 0.954 (0.951-0.957) | 0.917 (0.915-0.919) | 0.563 (0.561-0.565) |
| Over 18w after booster dose Adenovirus | 0.133 (0.112-0.158) | 0.149 (0.138-0.162) | 0.704 (0.685-0.724) | 0.867 (0.842-0.888) | 0.851 (0.838-0.862) | 0.296 (0.276-0.315) |
| Over 18w after booster dose mRNA | 0.056 (0.05-0.062) | 0.101 (0.097-0.106) | 0.696 (0.685-0.707) | 0.944 (0.938-0.95) | 0.899 (0.894-0.903) | 0.304 (0.293-0.315) |
| **WT** | | | | | | |
| Not Vaccinated | .. | 1 (1-1) | 1 (1-1) | .. | 0 (0-0) | 0 (0-0) |
| Less than 3w after first dose mRNA | .. | 0.169 (0.129-0.221) | 2.97 (2.497-3.533) | .. | 0.831 (0.779-0.871) | -1.97 (-2.533–1.497) |
| **Alpha** | | | | | | |
| Not Vaccinated | 1 (1-1) | 1 (1-1) | 1 (1-1) | 0 (0-0) | 0 (0-0) | 0 (0-0) |
| Less than 3w after first dose Adenovirus | 0.271 (0.26-0.283) | 0.675 (0.656-0.694) | 0.931 (0.92-0.943) | 0.729 (0.717-0.74) | 0.325 (0.306-0.344) | 0.069 (0.057-0.08) |
| Less than 3w after first dose mRNA | 0.139 (0.133-0.145) | 0.388 (0.378-0.399) | 0.856 (0.847-0.866) | 0.861 (0.855-0.867) | 0.612 (0.601-0.622) | 0.144 (0.134-0.153) |
| Over 3w after first dose Adenovirus | 0.326 (0.309-0.345) | 0.35 (0.335-0.366) | 0.521 (0.512-0.53) | 0.674 (0.655-0.691) | 0.65 (0.634-0.665) | 0.479 (0.47-0.488) |
| Over 3w after first dose mRNA | 0.136 (0.129-0.143) | 0.283 (0.273-0.294) | 0.564 (0.555-0.574) | 0.864 (0.857-0.871) | 0.717 (0.706-0.727) | 0.436 (0.426-0.445) |
| Less than 2w after second dose Adenovirus | .. | 0.195 (0.145-0.263) | 0.617 (0.563-0.675) | .. | 0.805 (0.737-0.855) | 0.383 (0.325-0.437) |
| Less than 2w after second dose mRNA | 0.031 (0.024-0.04) | 0.131 (0.116-0.148) | 0.321 (0.303-0.341) | 0.969 (0.96-0.976) | 0.869 (0.852-0.884) | 0.679 (0.659-0.697) |
| 2-10w after second dose Adenovirus | .. | 0.173 (0.121-0.248) | 0.622 (0.555-0.698) | .. | 0.827 (0.752-0.879) | 0.378 (0.302-0.445) |
| 2-10w after second dose mRNA | 0.023 (0.019-0.029) | 0.09 (0.079-0.102) | 0.231 (0.216-0.246) | 0.977 (0.971-0.981) | 0.91 (0.898-0.921) | 0.769 (0.754-0.784) |
| 10-18w after second dose mRNA | .. | 0.074 (0.043-0.128) | .. | .. | 0.926 (0.872-0.957) | .. |
| **Delta** | | | | | | |
| Not Vaccinated | 1 (1-1) | 1 (1-1) | 1 (1-1) | 0 (0-0) | 0 (0-0) | 0 (0-0) |
| Less than 3w after first dose Adenovirus | .. | 0.238 (0.193-0.292) | 0.511 (0.49-0.533) | .. | 0.762 (0.708-0.807) | 0.489 (0.467-0.51) |
| Less than 3w after first dose mRNA | 0.206 (0.128-0.333) | 0.319 (0.3-0.34) | 0.826 (0.822-0.831) | 0.794 (0.667-0.872) | 0.681 (0.66-0.7) | 0.174 (0.169-0.178) |
| Over 3w after first dose Adenovirus | 0.09 (0.079-0.103) | 0.096 (0.092-0.101) | 0.327 (0.325-0.33) | 0.91 (0.897-0.921) | 0.904 (0.899-0.908) | 0.673 (0.67-0.675) |
| Over 3w after first dose mRNA | 0.091 (0.078-0.108) | 0.131 (0.126-0.136) | 0.509 (0.507-0.511) | 0.909 (0.892-0.922) | 0.869 (0.864-0.874) | 0.491 (0.489-0.493) |
| Less than 2w after second dose Adenovirus | .. | 0.027 (0.023-0.03) | 0.283 (0.28-0.287) | .. | 0.973 (0.97-0.977) | 0.717 (0.713-0.72) |
| Less than 2w after second dose mRNA | .. | 0.048 (0.041-0.055) | 0.197 (0.195-0.199) | .. | 0.952 (0.945-0.959) | 0.803 (0.801-0.805) |
| 2-10w after second dose Adenovirus | 0.024 (0.02-0.028) | 0.022 (0.021-0.023) | 0.252 (0.25-0.253) | 0.976 (0.972-0.98) | 0.978 (0.977-0.979) | 0.748 (0.747-0.75) |
| 2-10w after second dose Mixed Dose | .. | .. | 0.223 (0.205-0.243) | .. | .. | 0.777 (0.757-0.795) |
| 2-10w after second dose mRNA | 0.016 (0.011-0.023) | 0.027 (0.025-0.029) | 0.098 (0.097-0.099) | 0.984 (0.977-0.989) | 0.973 (0.971-0.975) | 0.902 (0.901-0.903) |

continued on next page

continued from previous page

| Vaccine | IRR (Deaths) | IRR (Hospitalisations) | IRR (Pillar 2 PCR Cases) | Vaccine effectiveness (Deaths) | Vaccine effectiveness (Hospitalisations) | Vaccine effectiveness (Pillar 2 PCR Cases) |
|---|---|---|---|---|---|---|
| 10-18w after second dose Adenovirus | 0.042 (0.039-0.046) | 0.038 (0.036-0.039) | 0.323 (0.322-0.324) | 0.958 (0.954-0.961) | 0.962 (0.961-0.964) | 0.677 (0.676-0.678) |
| 10-18w after second dose Mixed Dose | .. | 0.102 (0.073-0.143) | 0.336 (0.315-0.359) | .. | 0.898 (0.857-0.927) | 0.664 (0.641-0.685) |
| 10-18w after second dose mRNA | 0.028 (0.024-0.031) | 0.027 (0.026-0.028) | 0.196 (0.195-0.197) | 0.972 (0.969-0.976) | 0.973 (0.972-0.974) | 0.804 (0.803-0.805) |
| Over 18w after second dose Adenovirus | 0.085 (0.081-0.09) | 0.066 (0.065-0.068) | 0.367 (0.365-0.368) | 0.915 (0.91-0.919) | 0.934 (0.932-0.935) | 0.633 (0.632-0.635) |
| Over 18w after second dose Mixed Dose | 0.174 (0.127-0.24) | 0.122 (0.101-0.148) | 0.412 (0.392-0.433) | 0.826 (0.76-0.873) | 0.878 (0.852-0.899) | 0.588 (0.567-0.608) |
| Over 18w after second dose mRNA | 0.069 (0.066-0.073) | 0.057 (0.056-0.059) | 0.296 (0.294-0.297) | 0.931 (0.927-0.934) | 0.943 (0.941-0.944) | 0.704 (0.703-0.706) |
| Less than 2w after booster dose Adenovirus | 0.012 (0.01-0.015) | 0.024 (0.022-0.025) | 0.208 (0.206-0.21) | 0.988 (0.985-0.99) | 0.976 (0.975-0.978) | 0.792 (0.79-0.794) |
| Less than 2w after booster dose mRNA | 0.014 (0.011-0.017) | 0.026 (0.024-0.027) | 0.167 (0.164-0.169) | 0.986 (0.983-0.989) | 0.974 (0.973-0.976) | 0.833 (0.831-0.836) |
| 2-10w after booster dose Adenovirus | 0.014 (0.012-0.017) | 0.014 (0.013-0.015) | 0.073 (0.072-0.075) | 0.986 (0.983-0.988) | 0.986 (0.985-0.987) | 0.927 (0.925-0.928) |
| 2-10w after booster dose mRNA | 0.012 (0.011-0.014) | 0.015 (0.014-0.016) | 0.092 (0.09-0.093) | 0.988 (0.986-0.989) | 0.985 (0.984-0.986) | 0.908 (0.907-0.91) |
| 10-18w after booster dose Adenovirus | .. | 0.033 (0.021-0.054) | 0.307 (0.283-0.332) | .. | 0.967 (0.946-0.979) | 0.693 (0.668-0.717) |
| 10-18w after booster dose mRNA | 0.009 (0.005-0.016) | 0.021 (0.017-0.025) | 0.158 (0.151-0.165) | 0.991 (0.984-0.995) | 0.979 (0.975-0.983) | 0.842 (0.835-0.849) |
| **Omicron** | | | | | | |
| Not Vaccinated | 1 (1-1) | 1 (1-1) | 1 (1-1) | 0 (0-0) | 0 (0-0) | 0 (0-0) |
| Less than 3w after first dose mRNA | .. | 0.5 (0.444-0.563) | 0.763 (0.752-0.773) | .. | 0.5 (0.437-0.556) | 0.237 (0.227-0.248) |
| Over 3w after first dose Adenovirus | 0.134 (0.11-0.163) | 0.492 (0.464-0.522) | 0.448 (0.44-0.455) | 0.866 (0.837-0.89) | 0.508 (0.478-0.536) | 0.552 (0.545-0.56) |
| Over 3w after first dose mRNA | 0.121 (0.098-0.149) | 0.295 (0.282-0.308) | 0.504 (0.501-0.507) | 0.879 (0.851-0.902) | 0.705 (0.692-0.718) | 0.496 (0.493-0.499) |
| Less than 2w after second dose mRNA | .. | 0.165 (0.14-0.194) | 0.35 (0.345-0.356) | .. | 0.835 (0.806-0.86) | 0.65 (0.644-0.655) |
| Less than 2w after second dose Mixed Dose | .. | 0.329 (0.23-0.471) | 0.43 (0.406-0.454) | .. | 0.671 (0.529-0.77) | 0.57 (0.546-0.594) |
| 2-10w after second dose Adenovirus | .. | 3.992 (2.912-5.472) | 10.257 (9.77-10.767) | .. | -2.992 (-4.472–1.912) | -9.257 (-9.767–8.77) |
| 2-10w after second dose Mixed Dose | 0.102 (0.058-0.18) | 0.175 (0.145-0.21) | 0.32 (0.309-0.331) | 0.898 (0.82-0.942) | 0.825 (0.79-0.855) | 0.68 (0.669-0.691) |
| 2-10w after second dose mRNA | .. | 0.213 (0.197-0.23) | 0.328 (0.325-0.331) | .. | 0.787 (0.77-0.803) | 0.672 (0.669-0.675) |
| 10-18w after second dose Adenovirus | .. | 5.788 (4.725-7.09) | 9.844 (9.521-10.178) | .. | -4.788 (-6.09–3.725) | -8.844 (-9.178–8.521) |
| 10-18w after second dose Mixed Dose | 0.313 (0.207-0.473) | 0.342 (0.275-0.426) | 0.689 (0.654-0.725) | 0.687 (0.527-0.793) | 0.658 (0.574-0.725) | 0.311 (0.275-0.346) |
| 10-18w after second dose mRNA | 0.093 (0.059-0.147) | 0.363 (0.343-0.384) | 0.65 (0.647-0.654) | 0.907 (0.853-0.941) | 0.637 (0.616-0.657) | 0.35 (0.346-0.353) |
| Over 18w after second dose Adenovirus | 0.299 (0.276-0.325) | 0.495 (0.48-0.51) | 0.657 (0.654-0.66) | 0.701 (0.675-0.724) | 0.505 (0.49-0.52) | 0.343 (0.34-0.346) |
| Over 18w after second dose Mixed Dose | 0.402 (0.255-0.632) | 0.439 (0.344-0.559) | 0.707 (0.674-0.742) | 0.598 (0.368-0.745) | 0.561 (0.441-0.656) | 0.293 (0.258-0.326) |
| Over 18w after second dose mRNA | 0.245 (0.223-0.27) | 0.361 (0.35-0.372) | 0.667 (0.665-0.67) | 0.755 (0.73-0.777) | 0.639 (0.628-0.65) | 0.333 (0.33-0.335) |
| Less than 2w after booster dose Adenovirus | 0.066 (0.052-0.085) | 0.136 (0.128-0.145) | 0.405 (0.403-0.408) | 0.934 (0.915-0.948) | 0.864 (0.855-0.872) | 0.595 (0.592-0.597) |
| Less than 2w after booster dose mRNA | 0.074 (0.048-0.114) | 0.16 (0.149-0.172) | 0.492 (0.489-0.495) | 0.926 (0.886-0.952) | 0.84 (0.828-0.851) | 0.508 (0.505-0.511) |
| 2-10w after booster dose Adenovirus | 0.042 (0.038-0.046) | 0.108 (0.104-0.111) | 0.305 (0.303-0.306) | 0.958 (0.954-0.962) | 0.892 (0.889-0.896) | 0.695 (0.694-0.697) |
| 2-10w after booster dose mRNA | 0.053 (0.047-0.059) | 0.152 (0.147-0.157) | 0.377 (0.375-0.379) | 0.947 (0.941-0.953) | 0.848 (0.843-0.853) | 0.623 (0.621-0.625) |
| 10-18w after booster dose Adenovirus | 0.066 (0.06-0.071) | 0.179 (0.174-0.185) | 0.45 (0.447-0.453) | 0.934 (0.929-0.94) | 0.821 (0.815-0.826) | 0.55 (0.547-0.553) |
| 10-18w after booster dose mRNA | 0.054 (0.05-0.058) | 0.173 (0.168-0.178) | 0.521 (0.518-0.524) | 0.946 (0.942-0.95) | 0.827 (0.822-0.832) | 0.479 (0.476-0.482) |
| Over 18w after booster dose Adenovirus | 0.124 (0.103-0.149) | 0.29 (0.267-0.315) | 0.83 (0.806-0.854) | 0.876 (0.851-0.897) | 0.71 (0.685-0.733) | 0.17 (0.146-0.194) |
| Over 18w after booster dose mRNA | 0.067 (0.059-0.075) | 0.215 (0.205-0.227) | 0.807 (0.794-0.819) | 0.933 (0.925-0.941) | 0.785 (0.773-0.795) | 0.193 (0.181-0.206) |

Table S3: Vaccine effectiveness estimates. Entries with .. represent estimates which have a p-value > 0.05.

| Ethnicity | IRR (Deaths) | IRR (Hospitalisations) | IRR (Pillar 2 PCR Cases) |
|---|---|---|---|
| **Full time period** | | | |
| White | 1 (1-1) | 1 (1-1) | 1 (1-1) |
| Black | 1.487 (1.432-1.543) | 1.555 (1.533-1.577) | 0.894 (0.892-0.897) |
| Mixed/Other | 0.895 (0.843-0.949) | 0.77 (0.756-0.785) | 0.429 (0.428-0.431) |
| Asian (other) | 1.242 (1.172-1.315) | 1.174 (1.15-1.199) | 0.784 (0.782-0.787) |
| South Asian | 1.633 (1.587-1.681) | 1.421 (1.404-1.438) | 0.885 (0.883-0.887) |
| **WT** | | | |
| White | 1 (1-1) | 1 (1-1) | 1 (1-1) |
| Black | 1.381 (1.28-1.489) | 1.64 (1.592-1.691) | 1.746 (1.689-1.804) |
| Mixed/Other | 0.749 (0.66-0.849) | 1.077 (1.036-1.12) | 1 (0.967-1.035) |
| Asian (other) | 1.322 (1.189-1.469) | 1.526 (1.468-1.587) | 1.557 (1.506-1.609) |
| South Asian | 1.801 (1.718-1.888) | 1.96 (1.92-2.002) | 2.883 (2.798-2.97) |
| **Alpha** | | | |
| White | 1 (1-1) | 1 (1-1) | 1 (1-1) |
| Black | 1.436 (1.362-1.514) | 1.87 (1.824-1.918) | 1.185 (1.178-1.193) |
| Mixed/Other | 0.93 (0.858-1.008) | 1.142 (1.104-1.18) | 0.664 (0.659-0.668) |
| Asian (other) | 1.323 (1.225-1.429) | 1.566 (1.514-1.62) | 1.035 (1.027-1.043) |
| South Asian | 1.634 (1.567-1.703) | 1.775 (1.738-1.813) | 1.444 (1.437-1.451) |
| **Delta** | | | |
| White | 1 (1-1) | 1 (1-1) | 1 (1-1) |
| Black | 1.45 (1.322-1.592) | 1.213 (1.181-1.247) | 0.735 (0.731-0.738) |
| Mixed/Other | 0.82 (0.706-0.954) | 0.58 (0.559-0.602) | 0.359 (0.357-0.361) |
| Asian (other) | 0.813 (0.681-0.971) | 0.795 (0.759-0.832) | 0.601 (0.597-0.605) |
| South Asian | 1.69 (1.567-1.823) | 1.143 (1.115-1.172) | 0.64 (0.637-0.642) |
| **Omicron** | | | |
| White | 1 (1-1) | 1 (1-1) | 1 (1-1) |
| Black | 0.989 (0.866-1.128) | 1.161 (1.125-1.198) | 0.9 (0.895-0.904) |
| Mixed/Other | 0.652 (0.527-0.806) | 0.482 (0.462-0.502) | 0.38 (0.377-0.382) |
| Asian (other) | 0.767 (0.604-0.973) | 0.744 (0.706-0.783) | 0.853 (0.848-0.859) |
| South Asian | 0.78 (0.684-0.888) | 0.85 (0.826-0.875) | 0.722 (0.719-0.726) |

Table S4: Ethnicity IRR estimates (no filter on p-values, all results displayed).

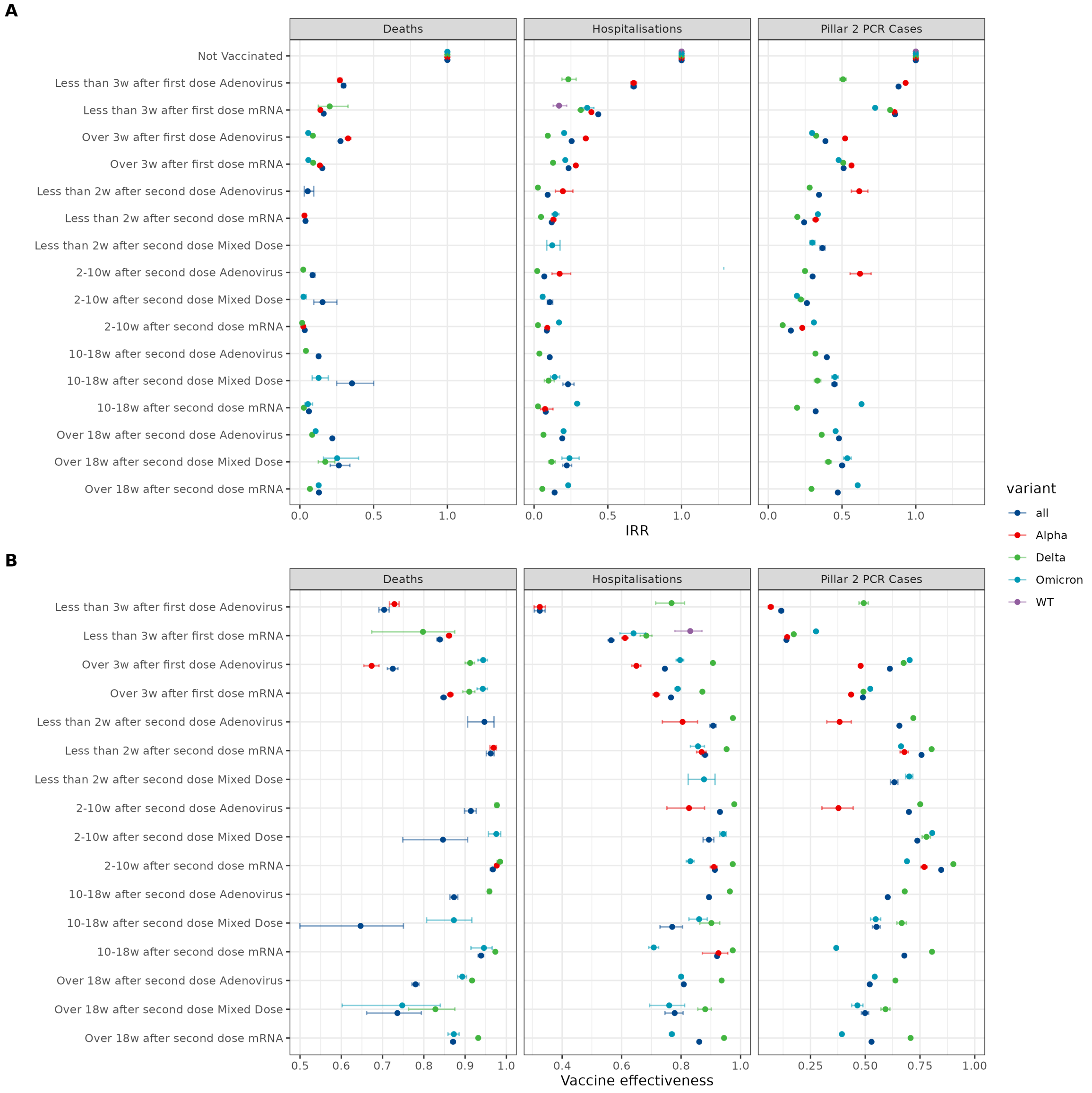

Figure S12: Vaccine effectiveness (VE=1-RR) with not vaccinated as the reference group (IRR=1 / VE=0), for different vaccination status. Results are shown for the whole pandemic ('all' - test dates between May 2020 and February 2022) and for the time intervals within that period where specific viral variants dominated (see SI for details). (A) IRR results, (B) Vaccine effectiveness results, WT=Wild-Type (pre-December 2020).

| Deprivation | IRR (Deaths) | IRR (Hospitalisations) | IRR (Pillar 2 PCR Cases) |
|---|---|---|---|
| **Full time period** | | | |
| IMD1 | 1 (1-1) | 1 (1-1) | 1 (1-1) |
| IMD2 | 1.084 (1.057-1.111) | 1.077 (1.063-1.09) | 0.978 (0.976-0.981) |
| IMD3 | 1.267 (1.237-1.297) | 1.179 (1.165-1.194) | 0.996 (0.994-0.998) |
| IMD4 | 1.366 (1.334-1.399) | 1.343 (1.327-1.36) | 1.007 (1.005-1.009) |
| IMD5 | 1.635 (1.596-1.674) | 1.527 (1.509-1.546) | 1.016 (1.014-1.018) |
| **WT** | | | |
| IMD1 | 1 (1-1) | 1 (1-1) | 1 (1-1) |
| IMD2 | 1.195 (1.144-1.247) | 1.183 (1.154-1.212) | 0.945 (0.911-0.98) |

continued on next page

continued from previous page

| Deprivation | IRR (Deaths) | IRR (Hospitalisations) | IRR (Pillar 2 PCR Cases) |
|---|---|---|---|
| IMD3 | 1.414 (1.356-1.474) | 1.37 (1.337-1.403) | 1.101 (1.061-1.142) |
| IMD4 | 1.553 (1.489-1.619) | 1.54 (1.504-1.577) | 1.189 (1.149-1.232) |
| IMD5 | 1.894 (1.819-1.973) | 1.851 (1.808-1.894) | 1.414 (1.366-1.465) |
| **Alpha** | | | |
| IMD1 | 1 (1-1) | 1 (1-1) | 1 (1-1) |
| IMD2 | 1.037 (1.002-1.073) | 1.093 (1.069-1.117) | 1.158 (1.152-1.165) |
| IMD3 | 1.24 (1.2-1.281) | 1.195 (1.17-1.22) | 1.284 (1.277-1.291) |
| IMD4 | 1.281 (1.239-1.325) | 1.361 (1.332-1.39) | 1.379 (1.371-1.386) |
| IMD5 | 1.501 (1.45-1.553) | 1.539 (1.507-1.573) | 1.453 (1.445-1.461) |
| **Delta** | | | |
| IMD1 | 1 (1-1) | 1 (1-1) | 1 (1-1) |
| IMD2 | 1.128 (1.047-1.215) | 1.056 (1.028-1.085) | 0.945 (0.942-0.948) |
| IMD3 | 1.376 (1.28-1.479) | 1.161 (1.131-1.191) | 0.934 (0.931-0.937) |
| IMD4 | 1.584 (1.475-1.701) | 1.319 (1.285-1.353) | 0.904 (0.901-0.907) |
| IMD5 | 1.895 (1.766-2.034) | 1.522 (1.484-1.56) | 0.866 (0.863-0.869) |
| **Omicron** | | | |
| IMD1 | 1 (1-1) | 1 (1-1) | 1 (1-1) |
| IMD2 | 1.014 (0.93-1.105) | 1.023 (0.996-1.052) | 0.981 (0.977-0.985) |
| IMD3 | 1.092 (1.004-1.188) | 1.046 (1.018-1.074) | 0.97 (0.966-0.974) |
| IMD4 | 1.274 (1.172-1.385) | 1.196 (1.165-1.228) | 0.962 (0.958-0.965) |
| IMD5 | 1.359 (1.249-1.478) | 1.22 (1.189-1.253) | 0.922 (0.919-0.926) |

Table S5: Deprivation (IMD national quintiles) IRR estimates (no filter on p-values, all results displayed).

#### D.1 IMD national deciles results

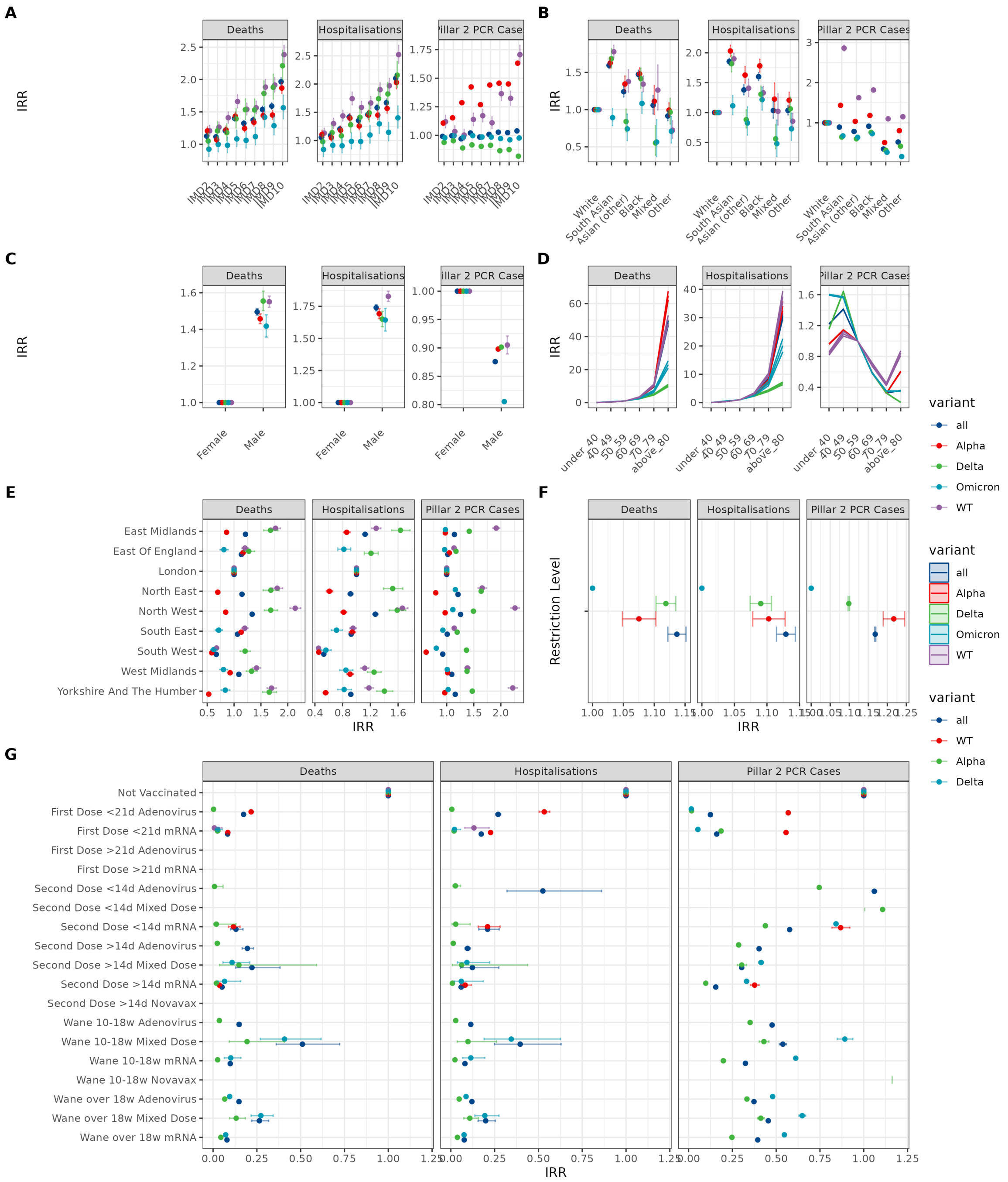

Figure S13: Results for the preferred model except that IMD national deciles are used as the deprivation measure. Estimated IRRs for (A) deprivation, (B) ethnicity, (C) sex, (D) age, (E) region, (F) restriction level, and (G) vaccine status covariates from preferred models. Deprivation was categorised by deciles of the Index of Multiple Deprivation score defined at LTLA level, with the first (least deprived) decile (IMD1) being the reference group (IRR=1). The reference group for age was 50-59 and for vaccine status reference group is not vaccinated. Results are shown for the whole pandemic ('all' - test dates between May 2020 and February 2022) and for the time intervals within that period where specific viral variants dominated (see SI for details). WT=Wild-Type (pre-December 2020).

#### D.2 Vaccine effectiveness comparison

Comparing vaccine effectiveness (VE) estimates is challenging due to differences in modelling choices, underlying data and time frames. In this section, we compare our estimates of VE (defined as 1-IRR) to those of Lopez-Bernal et al [8]. Lopez-Bernal et al used a test-negative case-control analysis and logistic regression methodology to estimate their model. The SARS-CoV-2 variant was determined at the individual level using the S-target negative/positive status and the authors ran the analysis on for individuals with the Alpha and Delta variant separately, allowing for a more granular comparison during periods when variants switch. In our analysis, we determine which is the majority variant (using S-target negative/positive status) for each week and region and variant represents a time period only. The time frame for this analysis was week 12 to 20 of 2021.

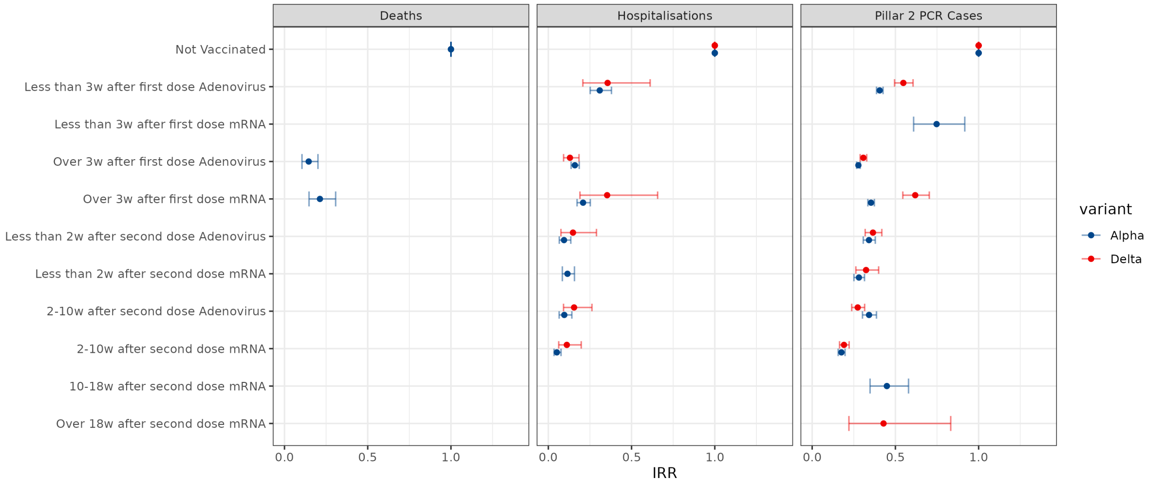

Figure S14: IRR estimates for vaccination status using the main model for weeks 12-20 (2021) consistent with the time frame for Lopez-Bernal et al [8].

Our methodology is less suited to picking up the difference in SARS-CoV-2 variants over short time periods, in particular when there is a switch in the dominant variant. All VE estimates below are with respect to Pillar 2 PCR-confirmed infections.

For the first dose, during week/region combinations, which we classified as Alpha, we observed a VE for mRNA vaccines of 64.6% (95% CI: 62.6-66.5) and for Delta a VE of 38.1% (95% CI: 29.6-45.5). This compares to the estimates for Lopez-Bernal et al of (48.7%; 95% CI, 45.5 to 51.7) for Alpha and (30.7%; 95% CI, 25.2 to 35.7) for Delta cases.

For the second dose of mRNA vaccines, we estimated a VE of 82.4% (95% CI:80.2-84.3) for the alpha period [Lopez-Bernal et al 93.7% (95% CI: 91.6-95.3)] and 80.9% (95% CI: 77.8-83.6) for the Delta period [Lopez-Bernal et al 88.0% (95% CI: 85.3-90.1)].

The estimates for the second dose of the Adenovirus vaccine were generally lower, in comparison to mRNA vaccines, with an estimated VE of 65.8% (95% CI: 61.4-69.8) for alpha [Lopez-Bernal et al of (74.5%; 95% CI: 68.4-79.4)] and 72.6% (95% CI: 68.4-76.2) for Delta [Lopez-Bernal et al of (67.0%; 95% CI: 61.3-71.8)].

If we consider the full Alpha and Delta periods, our results are consistent for Alpha but generally lower for Delta. We find VE for the mRNA vaccines ranged from 70.4% (second dose over 18 weeks) to 90.2% (second dose 2-10 weeks) versus 88.0% (95% CI, 85.3 to 90.1) for two doses of BNT162b2 for infections for persons with the delta variant. Similarly, we find vaccine effectiveness for the adenovirus vaccines ranged from 63.3% (second dose over 18 weeks) to 74.8% (second dose 2-10 weeks) compared to 67.0% (95% CI, 61.3 to 71.8) for persons with two doses of the ChAdOx1 nCoV-19 vaccine.

| Vaccine | IRR (Deaths) | IRR (Hospitalisations) | IRR (Pillar 2 PCR Cases) | Vaccine effectiveness (Deaths) | Vaccine effectiveness (Hospitalisations) | Vaccine effectiveness (Pillar 2 PCR Cases) |
|---|---|---|---|---|---|---|
| **Alpha** | | | | | | |
| Not Vaccinated | 1 (1-1) | 1 (1-1) | 1 (1-1) | 0 (0-0) | 0 (0-0) | 0 (0-0) |
| Less than 3w after first dose Adenovirus | .. | 0.309 (0.252-0.379) | 0.406 (0.387-0.426) | .. | 0.691 (0.621-0.748) | 0.594 (0.574-0.613) |
| Less than 3w after first dose mRNA | .. | .. | 0.747 (0.609-0.917) | .. | .. | 0.253 (0.083-0.391) |
| Over 3w after first dose Adenovirus | 0.145 (0.105-0.201) | 0.16 (0.137-0.186) | 0.278 (0.267-0.288) | 0.855 (0.799-0.895) | 0.84 (0.814-0.863) | 0.722 (0.712-0.733) |
| Over 3w after first dose mRNA | 0.213 (0.147-0.307) | 0.209 (0.173-0.252) | 0.354 (0.335-0.374) | 0.787 (0.693-0.853) | 0.791 (0.748-0.827) | 0.646 (0.626-0.665) |
| Less than 2w after second dose Adenovirus | .. | 0.094 (0.066-0.135) | 0.341 (0.307-0.379) | .. | 0.906 (0.865-0.934) | 0.659 (0.621-0.693) |
| Less than 2w after second dose mRNA | .. | 0.115 (0.084-0.157) | 0.281 (0.251-0.314) | .. | 0.885 (0.843-0.916) | 0.719 (0.686-0.749) |
| 2-10w after second dose Adenovirus | .. | 0.096 (0.065-0.141) | 0.342 (0.302-0.386) | .. | 0.904 (0.859-0.935) | 0.658 (0.614-0.698) |
| 2-10w after second dose mRNA | .. | 0.051 (0.034-0.076) | 0.176 (0.156-0.198) | .. | 0.949 (0.924-0.966) | 0.824 (0.802-0.844) |
| 10-18w after second dose mRNA | .. | .. | 0.449 (0.348-0.579) | .. | .. | 0.551 (0.421-0.652) |
| **Delta** | | | | | | |
| Not Vaccinated | .. | 1 (1-1) | 1 (1-1) | .. | 0 (0-0) | 0 (0-0) |
| Less than 3w after first dose Adenovirus | .. | 0.356 (0.207-0.612) | 0.548 (0.495-0.606) | .. | 0.644 (0.388-0.793) | 0.452 (0.394-0.505) |
| Over 3w after first dose Adenovirus | .. | 0.13 (0.092-0.184) | 0.307 (0.288-0.328) | .. | 0.87 (0.816-0.908) | 0.693 (0.672-0.712) |
| Over 3w after first dose mRNA | .. | 0.353 (0.19-0.657) | 0.619 (0.545-0.704) | .. | 0.647 (0.343-0.81) | 0.381 (0.296-0.455) |
| Less than 2w after second dose Adenovirus | .. | 0.148 (0.076-0.289) | 0.365 (0.319-0.418) | .. | 0.852 (0.711-0.924) | 0.635 (0.582-0.681) |
| Less than 2w after second dose mRNA | .. | .. | 0.324 (0.262-0.399) | .. | .. | 0.676 (0.601-0.738) |
| 2-10w after second dose Adenovirus | .. | 0.155 (0.091-0.262) | 0.274 (0.238-0.315) | .. | 0.845 (0.738-0.909) | 0.726 (0.685-0.762) |
| 2-10w after second dose mRNA | .. | 0.111 (0.062-0.198) | 0.191 (0.164-0.222) | .. | 0.889 (0.802-0.938) | 0.809 (0.778-0.836) |
| Over 18w after second dose mRNA | .. | .. | 0.429 (0.221-0.832) | .. | .. | 0.571 (0.168-0.779) |

Table S6: Vaccine effectiveness estimates using the main model for weeks 12-20 (2021) consistent with the time frame for Lopez-Bernal et al [8].

#### D.3 Regional resolution sensitivity analysis

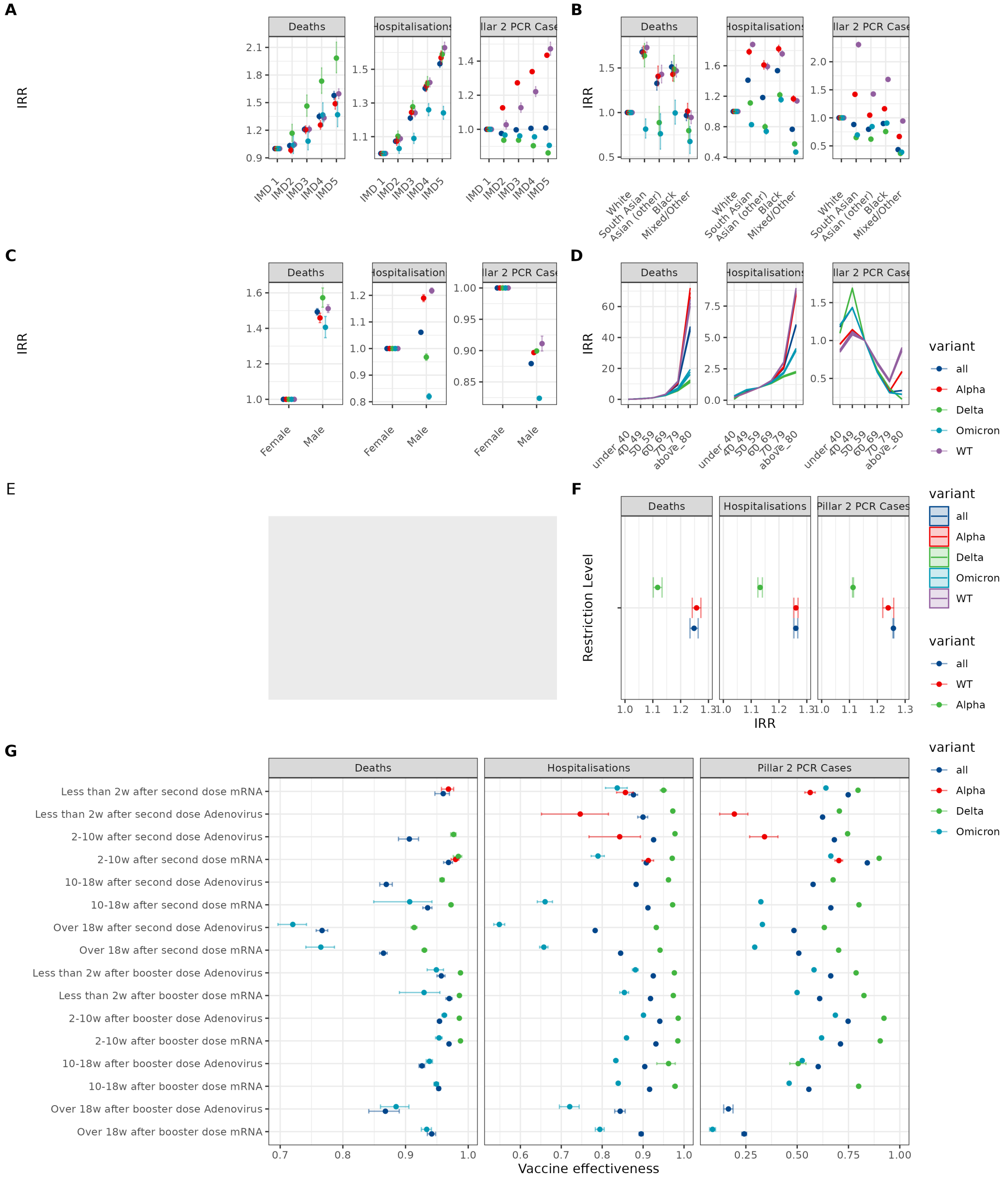

Figure S15: Results for the preferred model except that ITL221 regions are used as the region id. Estimated IRRs for (A) deprivation, (B) ethnicity, (C) sex, (D) age, (E) too many regions to display (33), (F) restriction level, and (G) vaccine status covariates from preferred models. Deprivation was categorised by quintiles of the IMD score defined at LTLA level, with the first (least deprived) decile (IMD 1) being the reference group (IRR=1). The reference group for age was 50-59 and for vaccine status reference group is not vaccinated. Results are shown for the whole pandemic ('all' - test dates between May 2020 and February 2022) and for the time intervals within that period where specific viral variants dominated (see SI for details). WT=Wild-Type (pre-December 2020).

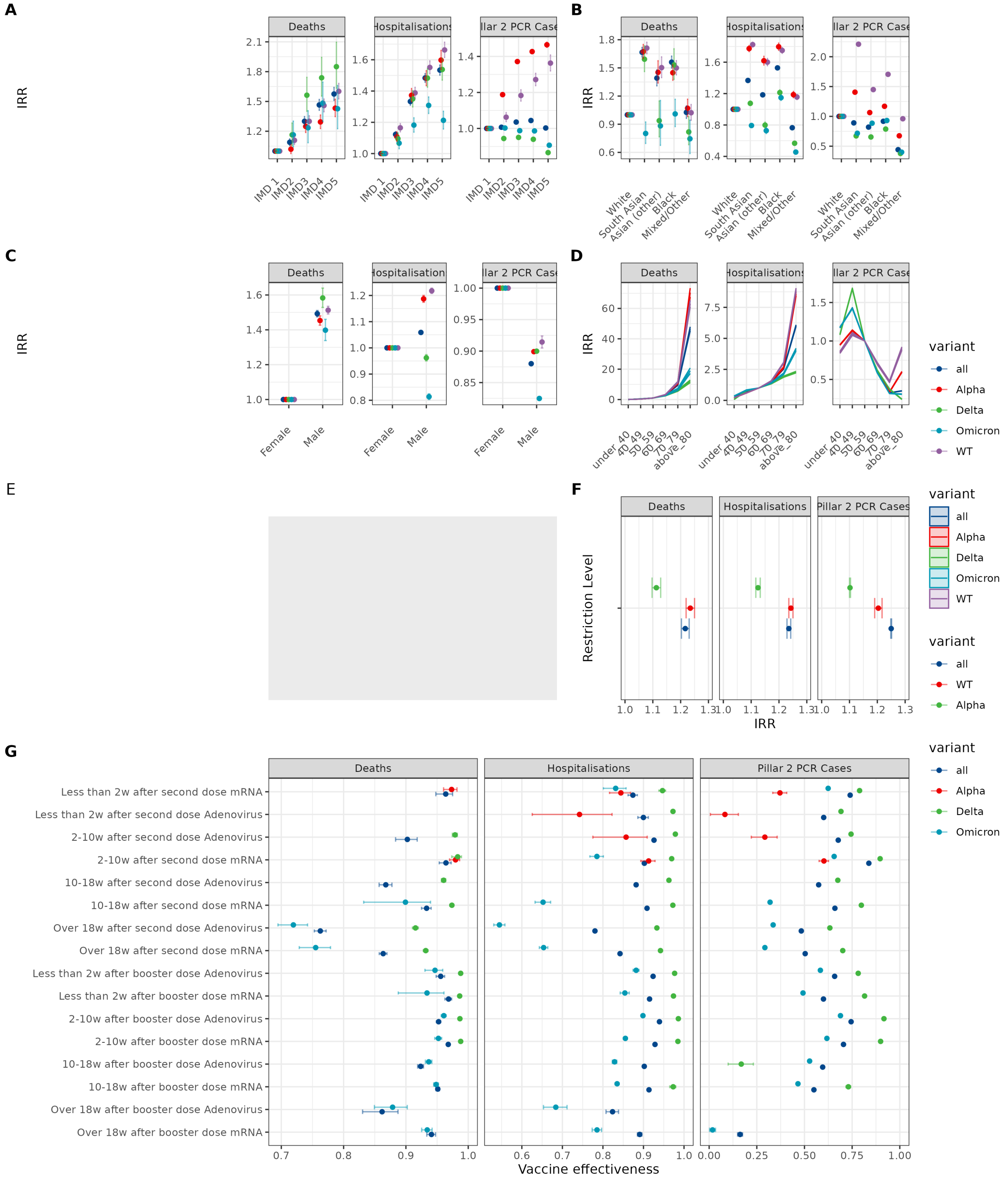

Figure S16: Results for the preferred model except that IRL321 regions are used as the region id. Estimated IRRs for (A) deprivation, (B) ethnicity, (C) sex, (D) age, (E) too many regions to display (133), (F) restriction level, and (G) vaccine status covariates from preferred models. Deprivation was categorised by quintiles of the IMD score defined at LTLA level, with the first (least deprived) decile (IMD 1) being the reference group (IRR=1). The reference group for age was 50-59 and for vaccine status reference group is not vaccinated. Results are shown for the whole pandemic ('all' - test dates between May 2020 and February 2022) and for the time intervals within that period where specific viral variants dominated (see SI for details). WT=Wild-Type (pre-December 2020).

#### D.4 Vaccine Effectiveness interactions

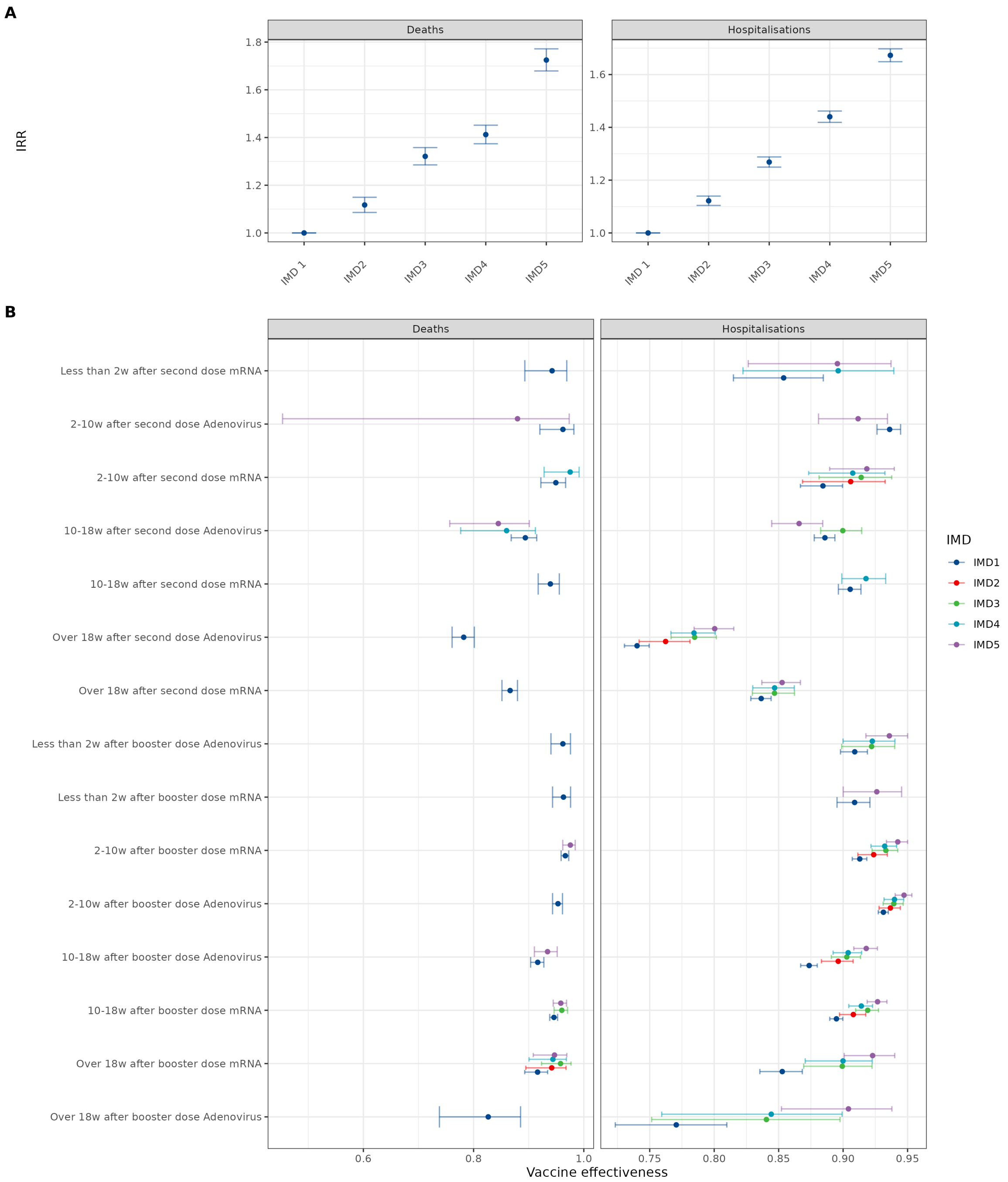

Figure S17: Results for the preferred model including an interaction term for vaccine status and IMD. Estimated IRRs for (A) deprivation, (B) vaccine effectiveness interaction with IMD. Deprivation was categorised by quintiles of the IMD score defined at LTLA level, with the first (least deprived) decile (IMD 1) being the reference group (IRR=1). The reference group for vaccine status reference group is not vaccinated and the interaction term vaccine status x IMD1. Results are shown for the whole pandemic ('all' - test dates between May 2020 and February 2022).

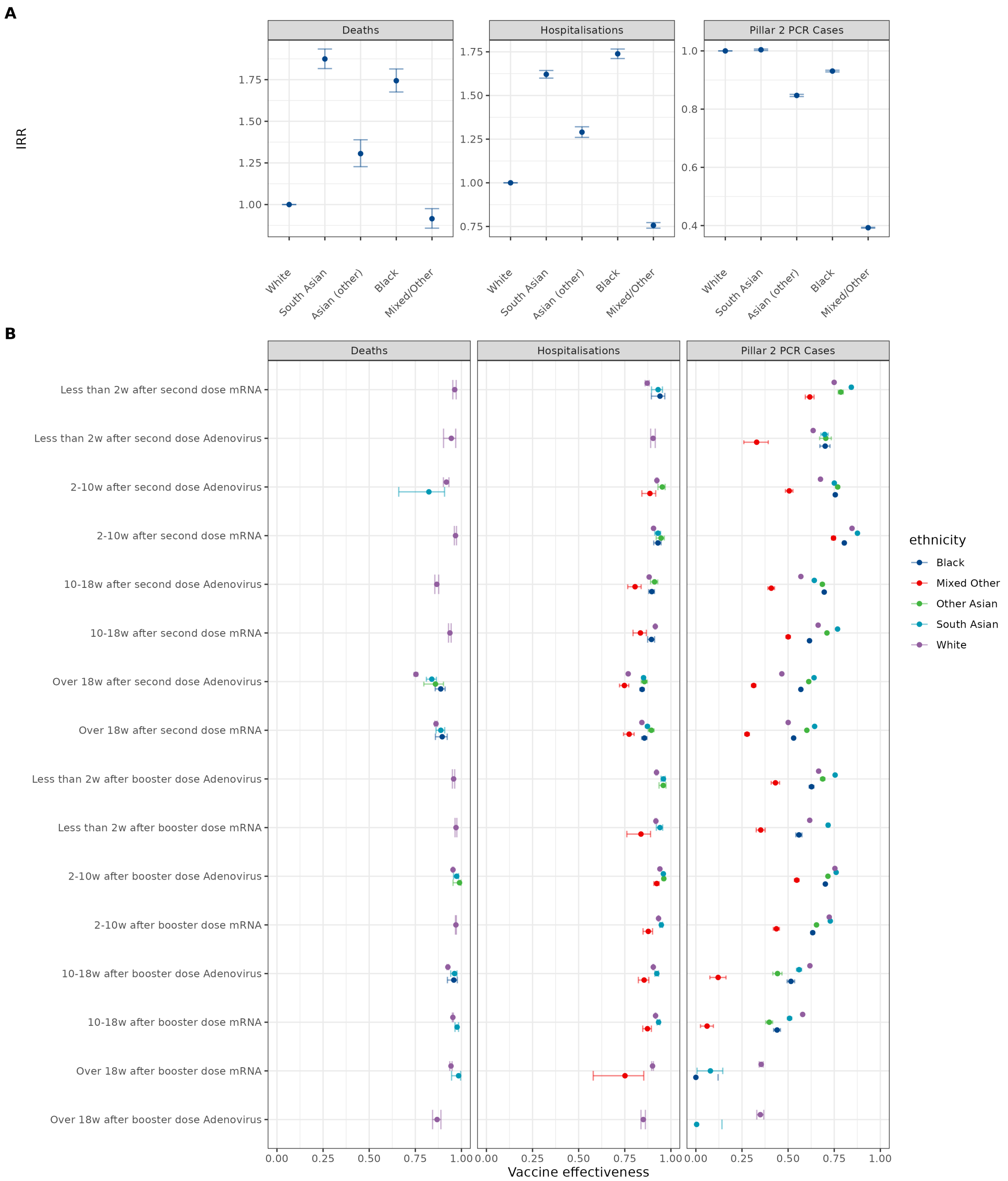

Figure S18: Results for the preferred model including an interaction term for vaccine status and ethnicity. Estimated IRRs for (A) Ethnicity, (B) vaccine effectiveness interaction with ethnicity. White ethnicity is the reference group (IRR=1). The reference group for vaccine status reference group is not vaccinated and the interaction term vaccine status x White ethnicity. Results are shown for the whole pandemic ('all' - test dates between May 2020 and February 2022).

### E Index of Multiple Deprivation Subcomponent Analysis

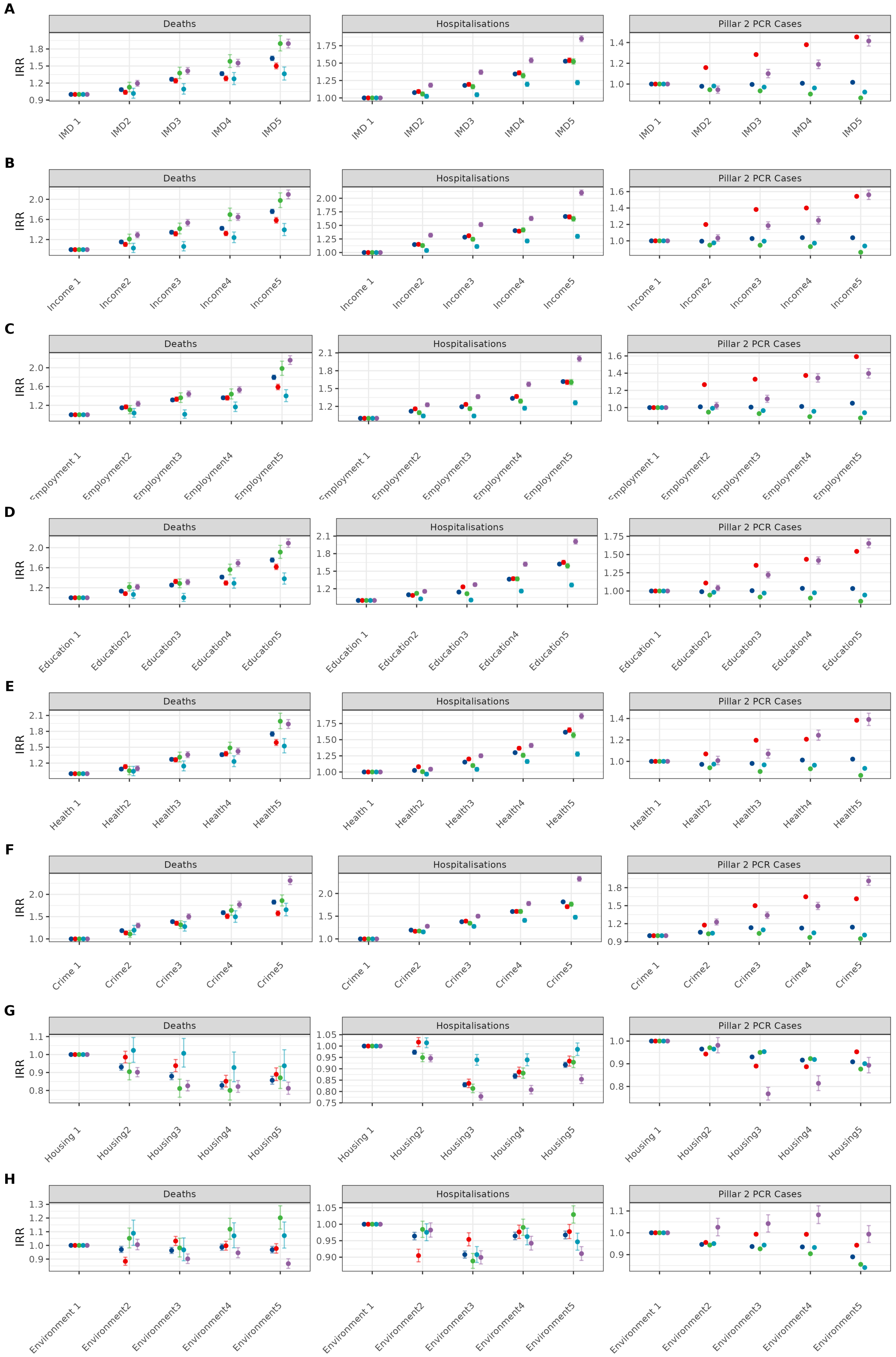

Figure S19: Index of Multiple Deprivation: Overall index (quintiles) and the 7 sub-components. Note that Housing is the inverse score, that is Housing 1 is the least deprived and Housing 5 the most deprived.

#### E.1 Income quintiles

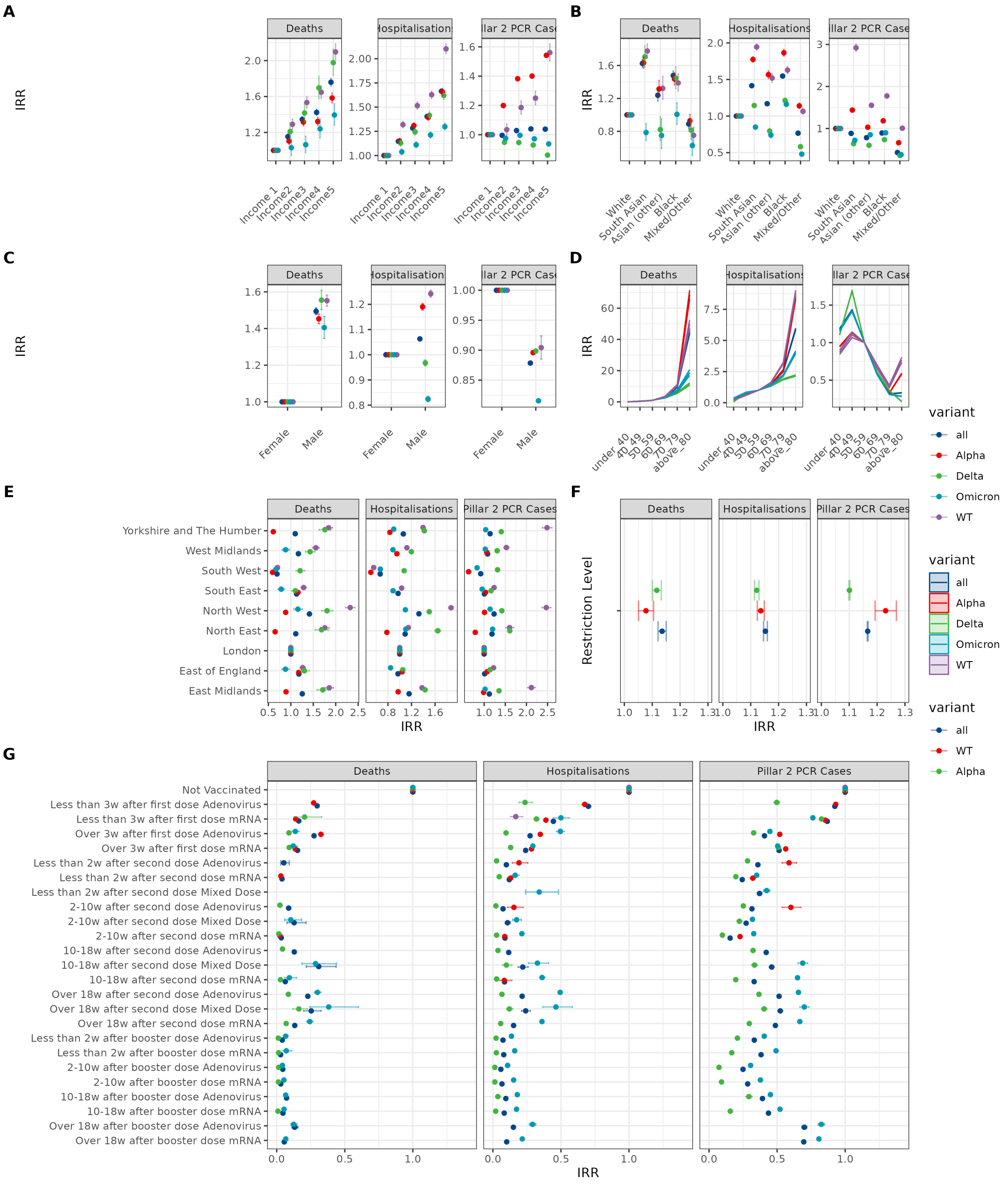

Figure S20: Results for the preferred model except that Income quintiles are used as the deprivation measure. Estimated IRRs for (A) deprivation, (B) ethnicity, (C) sex, (D) age, (E) region, (F) restriction level, and (G) vaccine status covariates from preferred models. Deprivation was categorised by quintiles of the Income Deprivation score defined at LTLA level, with the first (least deprived) decile (Income 1) being the reference group (IRR=1). The reference group for age was 50-59 and for vaccine status reference group is not vaccinated. Results are shown for the whole pandemic ('all' - test dates between May 2020 and February 2022) and for the time intervals within that period where specific viral variants dominated (see SI for details). WT=Wild-Type (pre-December 2020).

#### E.2 Employment quintiles

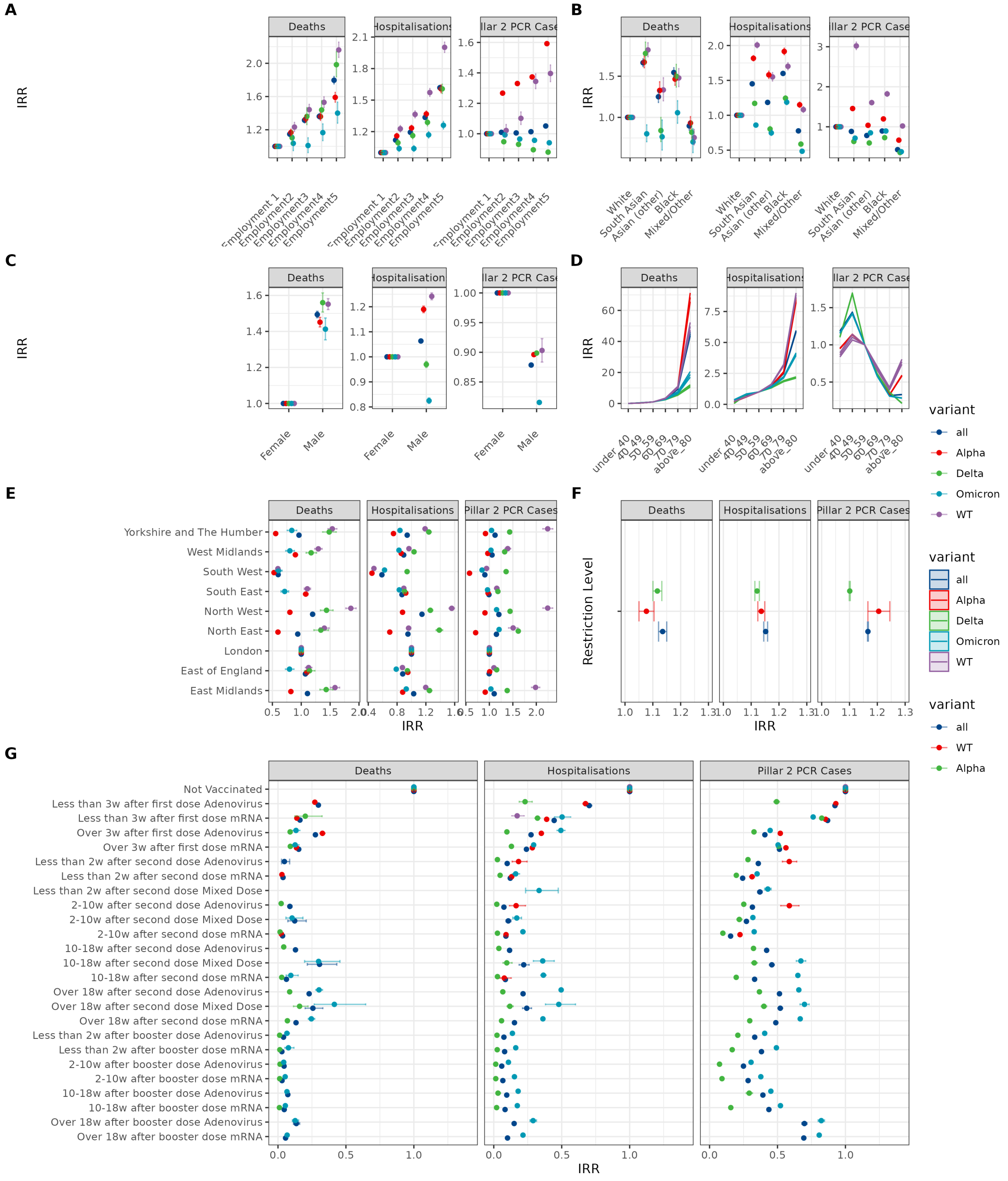

Figure S21: Results for the preferred model except that Employment quintiles are used as the deprivation measure. Estimated IRRs for (A) deprivation, (B) ethnicity, (C) sex, (D) age, (E) region, (F) restriction level, and (G) vaccine status covariates from preferred models. Deprivation was categorised by quintiles of the Employment Deprivation score defined at LTLA level, with the first (least deprived) decile (Employment 1) being the reference group (IRR=1). The reference group for age was 50-59 and for vaccine status reference group is not vaccinated. Results are shown for the whole pandemic ('all' - test dates between May 2020 and February 2022) and for the time intervals within that period where specific viral variants dominated (see SI for details). WT=Wild-Type (pre-December 2020).

#### E.3 Education quintiles

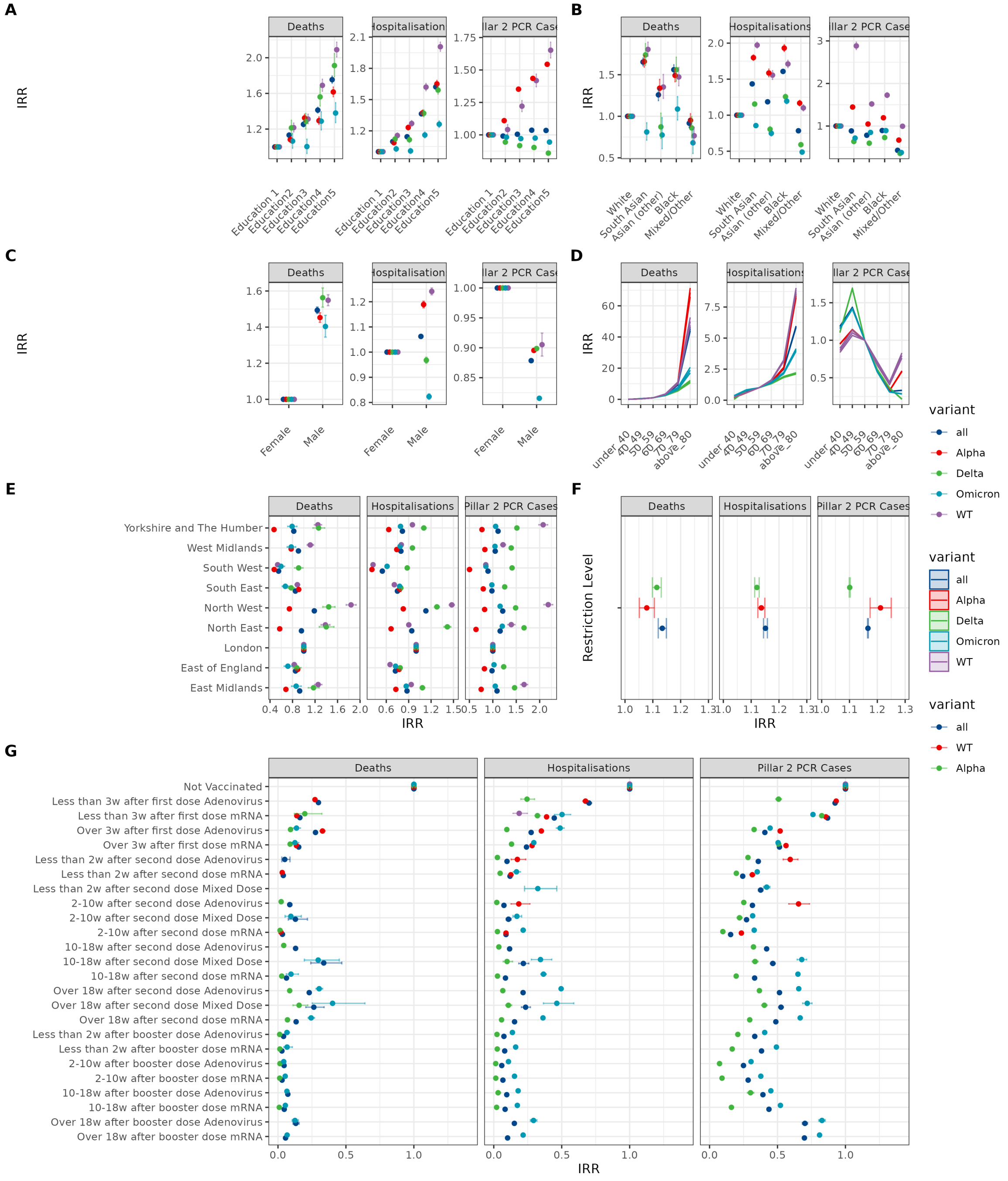

Figure S22: Results for the preferred model except that Education quintiles are used as the deprivation measure. Estimated IRRs for (A) deprivation, (B) ethnicity, (C) sex, (D) age, (E) region, (F) restriction level, and (G) vaccine status covariates from preferred models. Deprivation was categorised by quintiles of the Education Deprivation score defined at LTLA level, with the first (least deprived) decile (Education 1) being the reference group (IRR=1). The reference group for age was 50-59 and for vaccine status reference group is not vaccinated. Results are shown for the whole pandemic ('all' - test dates between May 2020 and February 2022) and for the time intervals within that period where specific viral variants dominated (see SI for details). WT=Wild-Type (pre-December 2020).

#### E.4 Health quintiles

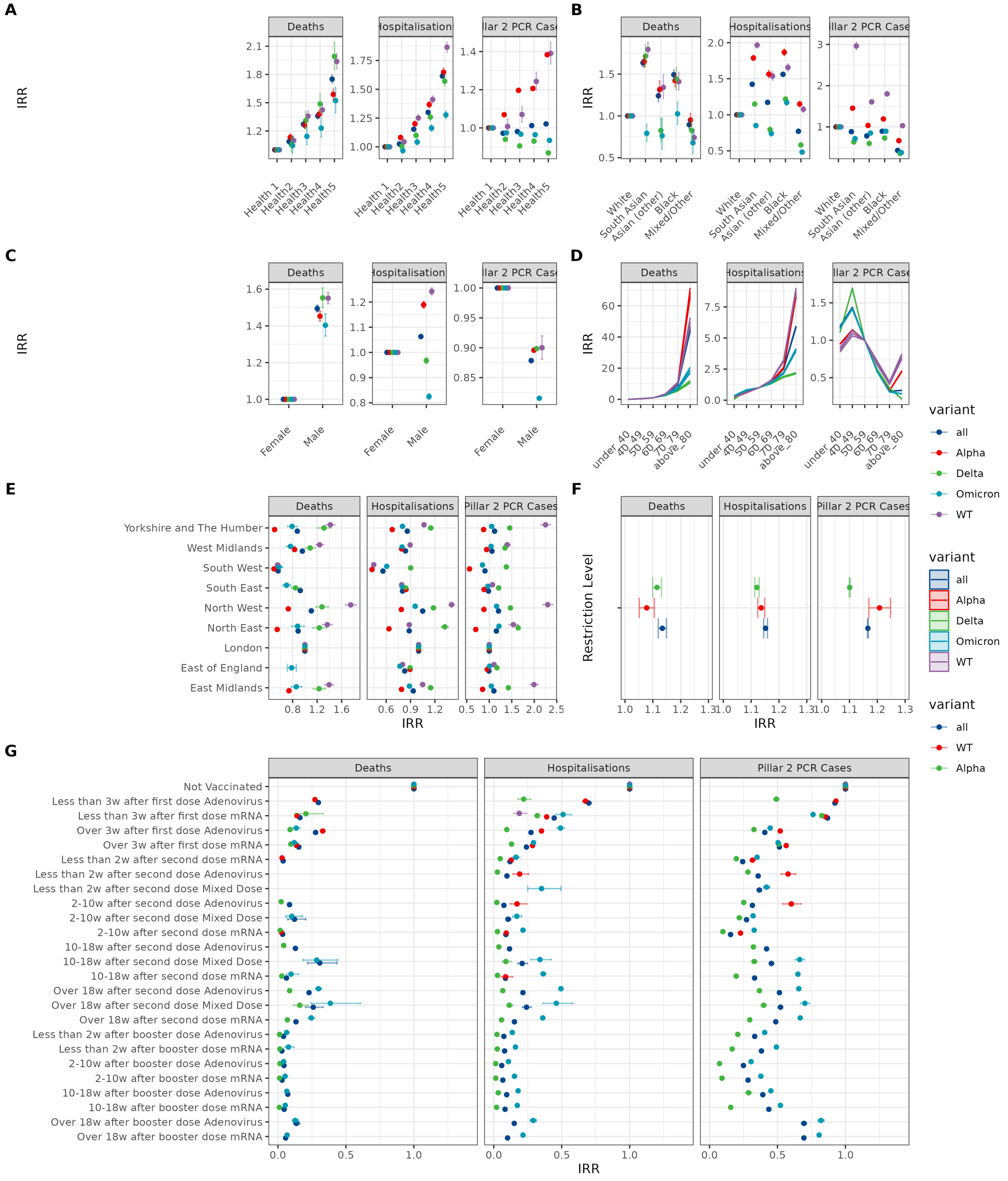

Figure S23: Results for the preferred model except that Health quintiles are used as the deprivation measure. Estimated IRRs for (A) deprivation, (B) ethnicity, (C) sex, (D) age, (E) region, (F) restriction level, and (G) vaccine status covariates from preferred models. Deprivation was categorised by quintiles of the Health Deprivation score defined at LTLA level, with the first (least deprived) decile (Health 1) being the reference group (IRR=1). The reference group for age was 50-59 and for vaccine status reference group is not vaccinated. Results are shown for the whole pandemic ('all' - test dates between May 2020 and February 2022) and for the time intervals within that period where specific viral variants dominated (see SI for details). WT=Wild-Type (pre-December 2020).

#### E.5 Crime quintiles

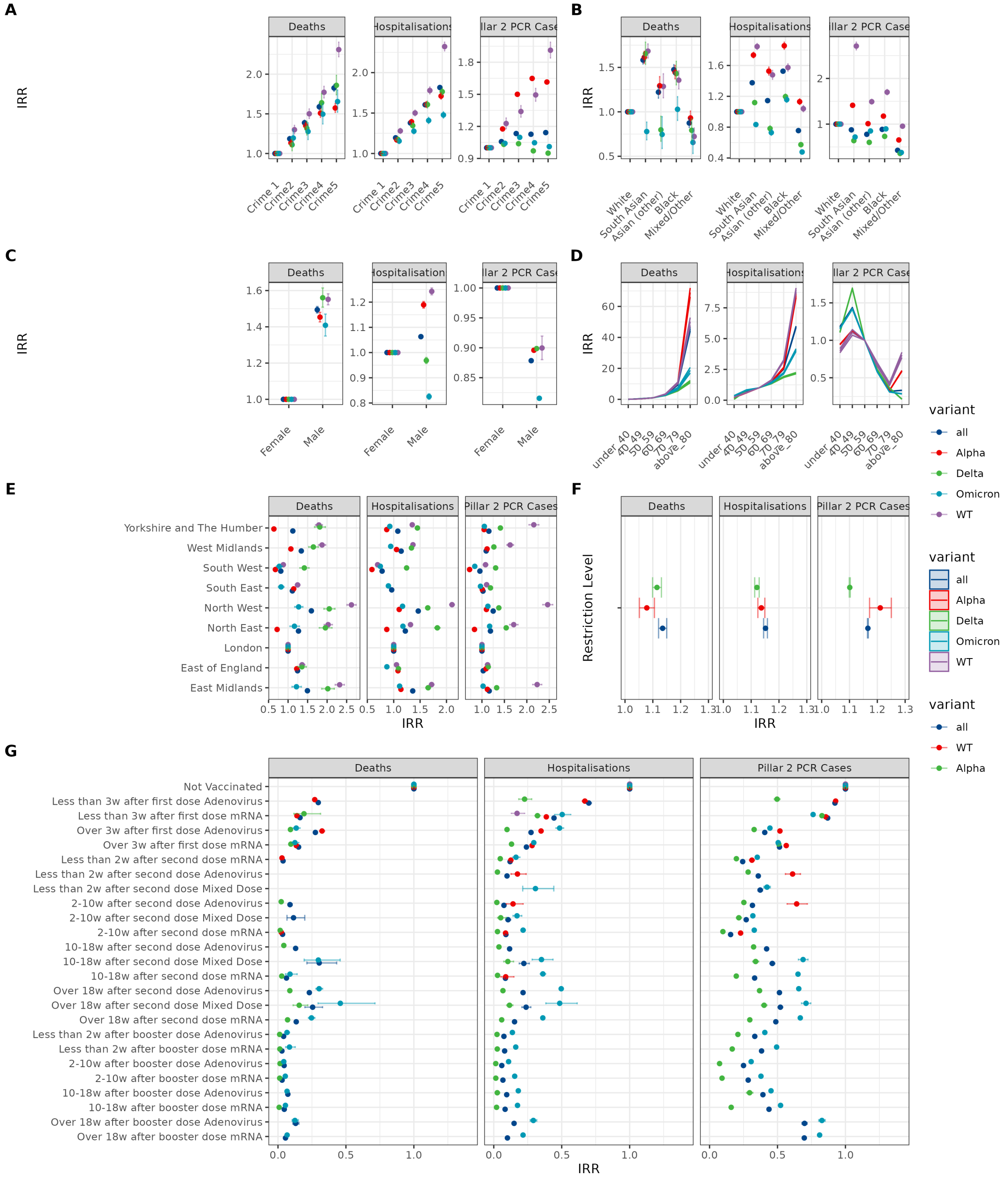

Figure S24: Results for the preferred model except that Crime quintiles are used as the deprivation measure. Estimated IRRs for (A) deprivation, (B) ethnicity, (C) sex, (D) age, (E) region, (F) restriction level, and (G) vaccine status covariates from preferred models. Deprivation was categorised by quintiles of the Crime Deprivation score defined at LTLA level, with the first (least deprived) decile (Crime 1) being the reference group (IRR=1). The reference group for age was 50-59 and for vaccine status reference group is not vaccinated. Results are shown for the whole pandemic ('all' - test dates between May 2020 and February 2022) and for the time intervals within that period where specific viral variants dominated (see SI for details). WT=Wild-Type (pre-December 2020).

#### E.6 Housing quintiles

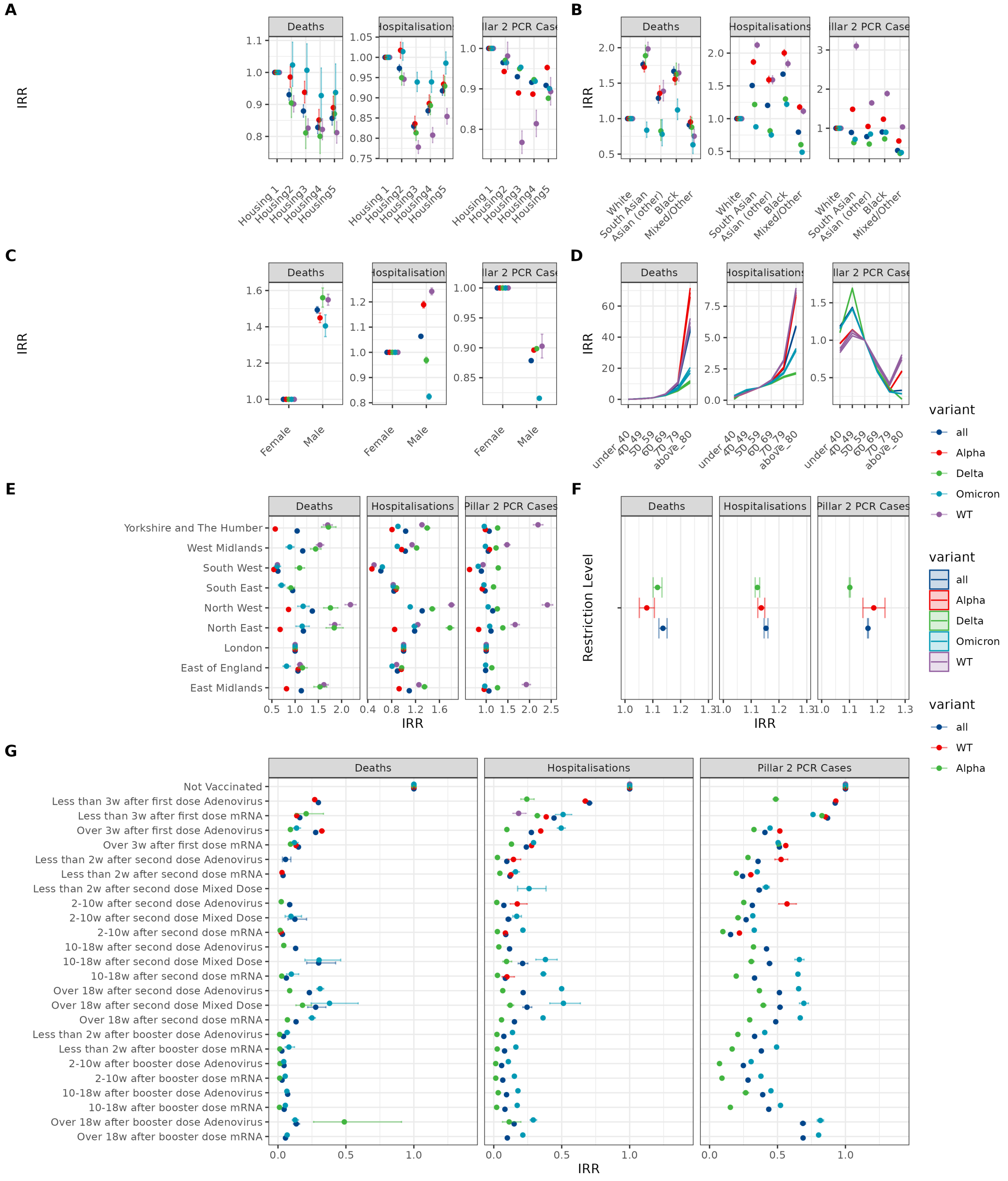

Figure S25: Results for the preferred model except that Housing quintiles are used as the deprivation measure. Estimated IRRs for (A) deprivation, (B) ethnicity, (C) sex, (D) age, (E) region, (F) restriction level, and (G) vaccine status covariates from preferred models. Deprivation was categorised by quintiles of the Housing Deprivation score defined at LTLA level, with the first (most deprived) decile (Housing 1) being the reference group (IRR=1). The reference group for age was 50-59 and for vaccine status reference group is not vaccinated. Results are shown for the whole pandemic ('all' - test dates between May 2020 and February 2022) and for the time intervals within that period where specific viral variants dominated (see SI for details). WT=Wild-Type (pre-December 2020).

#### E.7 Environmental quintiles

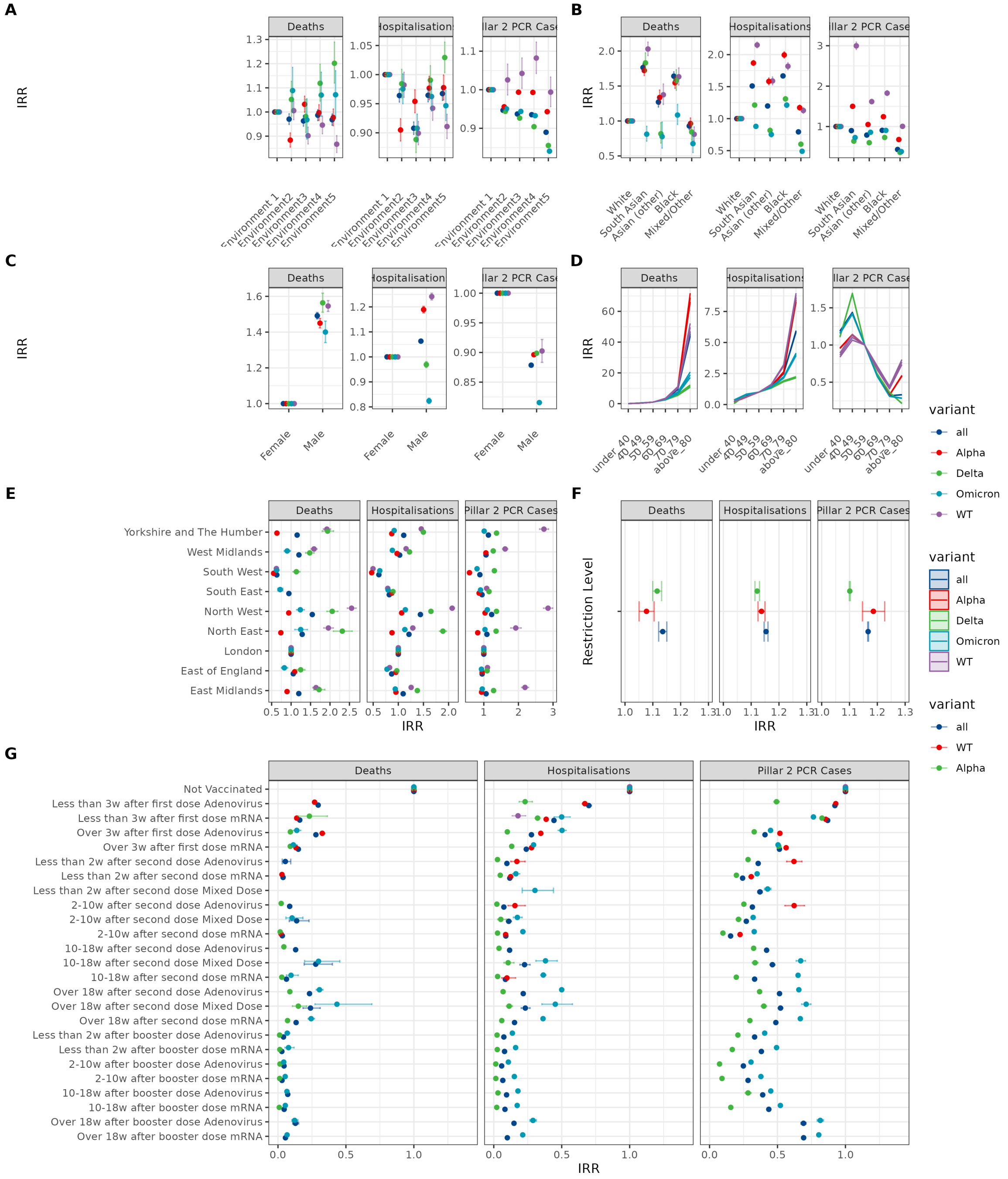

Figure S26: Results for the preferred model except that Environment quintiles are used as the deprivation measure. Estimated IRRs for (A) deprivation, (B) ethnicity, (C) sex, (D) age, (E) region, (F) restriction level, and (G) vaccine status covariates from preferred models. Deprivation was categorised by quintiles of the Environment Deprivation score defined at LTLA level, with the first (least deprived) decile (Environment 1) being the reference group (IRR=1). The reference group for age was 50-59 and for vaccine status reference group is not vaccinated. Results are shown for the whole pandemic ('all' - test dates between May 2020 and February 2022) and for the time intervals within that period where specific viral variants dominated (see SI for details). WT=Wild-Type (pre-December 2020).
